# Decomposing cognitive-motor planning from execution: smartphone motor sequencing provides scalable digital biomarkers of cognitive-motor function across central nervous system disorders

**DOI:** 10.64898/2026.07.30.26359238

**Authors:** Peter Kosa, Amir Moghadam Ahmadi, Marie Kanu, Yolanda Mejia, Emmanuel Mekasha, Caledonia Steltzner, Bibiana Bielekova

## Abstract

Dynamic praxis, defined as the ability to plan and execute complex, ordered motor actions, underpins essential activities of daily living and occupational performance. Because motor sequencing depends on distributed frontostriatal and interhemispheric networks, its impairment serves as a sensitive indicator of central nervous system (CNS) dysfunction, yet traditional bedside assessments lack granular subprocess resolution. Here, we digitized Luria’s fist-edge-palm paradigm into a self-administered smartphone task within the Neurological Functional Test Suite (NeuFun-TS) and evaluated its clinical validity in 296 participants (34 healthy donors, 208 people with multiple sclerosis (MS), and 54 neurological controls). We extracted seven digital biomarkers across speed, execution, accuracy, and integrative throughput. Six biomarkers significantly differentiated disease cohorts along an ordinal severity gradient (healthy donors < relapsing-remitting MS < progressive MS), with Motor Sequencing Deficit showing the strongest group separation (r=0.73, p=1.1×10^-21^). Kinematic decomposition dissociated cognitive planning latency (ε^2^=0.011) from pure motor execution (ε ^2^=0.716). Speed-accuracy tradeoff analysis differentiated secondary-progressive MS (84% slow-and-inaccurate) from primary-progressive MS (19% slow-but-accurate compensatory phenotype). A parsimonious two-biomarker composite achieved high diagnostic classification accuracy (concordance index = 0.868; validation intraclass correlation coefficient ICC = 0.77) and correlated strongly with CNS tissue destruction on MRI (rho=0.29-0.38), clinician disability scales (rho=0.44-0.52), and cognitive performance (rho=0.49-0.61). By capturing subtle cognitive-motor planning and execution deficits, smartphone-based motor sequencing offers a scalable, low-burden framework for longitudinal neurological monitoring in MS and broader central nervous system disorders affecting daily functional independence.

## Introduction

The ability to plan, maintain, and execute an ordered series of distinct motor actions into a seamless, automated flow represents a critical higher cortical function termed dynamic praxis or motor sequencing. This capability underpins a vast array of skilled human activities and daily routines, including manual manufacturing assembly, precision surgical procedures, and fine arts and crafts. Consequently, deficits in motor sequencing are predicted to adversely affect occupational performance and impair an individual’s independence in daily living functions.

In his seminal monograph Higher Cortical Functions in Man, Alexander Luria demonstrated that motor sequencing depends on distributed frontal-subcortical networks rather than the primary motor cortex alone (Luria, 1966). Luria’s fist-edge-palm task (FEP), in which individuals must reproduce a three-position sequential hand posture on a flat surface, remains one of the most widely used bedside assessments of dynamic praxis and was formalized as the “Motor Series” subtest of the Frontal Assessment Battery (Dubois et al., 2000). Unlike traditional neurological tests evaluating isolated motor strength or localized manual dexterity, the FEP task isolates the temporal organization and chronological serialization of movements. Modern neuroanatomy has localized these sequential functions to a network comprising the supplementary motor area (SMA), pre-SMA, basal ganglia, and prefrontal cortex (Tanji, 2001). Functional magnetic resonance imaging (fMRI) has confirmed that the FEP task (with either hand) engages significantly more extensive cortical and subcortical networks than simple repetitive movements, specifically eliciting activation within bilateral premotor areas, the left parietal cortex, the ipsilateral cerebellum as well as contralateral sensory-motor area and SMA (Umetsu et al., 2002). Furthermore, FEP and many other motor sequencing tasks must also engage cognitive mechanisms associated with the inhibition of the dominant but incorrect movements, and this inhibitory role was localized to the right prefrontal cortex (Varkovetski et al., 2020).

The distributed network underlying motor sequencing is uniquely vulnerable to multiple sclerosis (MS) pathology: because motor programming relies on long-range white matter tracts connecting bilateral cortical regions with subcortical nuclei and the cerebellum, this circuitry is highly susceptible to disruption by focal inflammatory and demyelinating lesions. The corpus callosum, which facilitates the interhemispheric coordination, is among the earliest and most severely affected white matter structures in MS (Bodini et al., 2013). Its microstructural integrity, measured via diffusion tensor imaging fractional anisotropy, correlates highly (r = 0.93) with explicit visuomotor sequence learning, even in people with MS (pwMS) who present with minimal clinical disability (Bonzano et al., 2011). Furthermore, the SMA interhemispheric tract, which is critical for coordinating concurrent motor and cognitive demands, exhibits specific diffusivity alterations in pwMS that directly predict motor variability during dual-task execution (Fritz et al., 2019).

While preliminary observations from our group (B.B.) indicate that pwMS often require more extensive repetition than healthy individuals to perform the FEP task flawlessly, especially within a timed paradigm, the traditional ordinal scoring of an untimed bedside FEP task failed to identify significant differences between 30 pwMS and 33 neurologically intact controls (Beatty & Monson, 1994). Consequently, we hypothesized that a more sensitive, digitally augmented motor sequencing assessment could identify and longitudinally track demyelination-associated slowing and/or deficits in pwMS and thus expand evaluation of higher cognitive functions in daily clinical practice.

In this paper we describe development of motor sequencing test amenable to convenient, self-administration via smartphone, its automated scoring, extraction of relevant digital biomarkers and assessment of their clinical value in pwMS.

## Results

### Cohort description and task implementation

Across all cohorts, 296 unique participants completed 3,278 motor sequencing trials (Table 1; Supplementary Figure S1). The full cohort comprised healthy donors (HD, n = 34 participants), relapsing-remitting MS (RR-MS, n = 101), secondary-progressive MS (SP-MS, n = 55), primary-progressive MS (PP-MS, n = 52), non-inflammatory neurological disease (NIND, n = 25), other inflammatory neurological disease (OIND, n = 21), and clinically or radiologically isolated syndrome (CIS/RIS, n = 8). We limited all primary analyses to HD and MS participants (241 participants, 479 first-trial patient-hand observations: HD = 65 hands from 33 participants, RR-MS = 202 from 101, SP-MS = 110 from 55, PP-MS = 102 from 52), reserving the non-MS diagnostic groups as an independent test of discriminant validity.

**Table 1.** Study population demographics at first motor sequencing assessment.

| Group | N | Age (years) | Female,<br>n (%) | Disease<br>duration (years) | EDSS | Trials (L/R) |
| --- | --- | --- | --- | --- | --- | --- |
| HD <sup>(a)</sup> | 34 | 37.4<br>[24.8, 52.1] <sup>c,d</sup> | 17/33<br>(52%) | NA | 1.8<br>[1.1, 2.8] <sup>b,c,d,f</sup> | 188 / 183 |
| RR-MS <sup>(b)</sup> | 101 | 49.3<br>[38.3, 56.9] <sup>c,d</sup> | 67/101<br>(66%) | 13.0<br>[5.2, 23.6] <sup>c</sup> | 3.5<br>[2.5, 5.0] <sup>a,c,d</sup> | 450 / 442 |
| SP-MS <sup>(c)</sup> | 55 | 60.7<br>[50.9, 66.8] <sup>a,b,e,f</sup> | 36/55<br>(65%) | 22.1<br>[14.2, 33.2] <sup>b,e,f</sup> | 6.0<br>[5.0, 6.5] <sup>a,b,e,f</sup> | 653 / 648 |
| PP-MS <sup>(d)</sup> | 52 | 63.1<br>[56.4, 68.3] <sup>a,b,e,f</sup> | 28/52<br>(54%) | 14.0<br>[11.5, 24.6] <sup>e,f</sup> | 6.0<br>[5.0, 6.5] <sup>a,b,e,f</sup> | 260 / 237 |
| NIND <sup>(e)</sup> | 25 | 47.3<br>[34.7, 59.7] <sup>c,d</sup> | 19/25<br>(76%) | 5.8<br>[2.6, 13.7] <sup>c,d</sup> | 3.5<br>[2.0, 4.5] <sup>c,d</sup> | 37 / 37 |
| OIND <sup>(f)</sup> | 21 | 42.1<br>[30.5, 49.6] <sup>c,d</sup> | 11/21<br>(52%) | 6.9<br>[2.8, 11.0] <sup>c,d</sup> | 4.0<br>[2.5, 6.0] <sup>a,c,d</sup> | 34 / 34 |
| CIS/RIS <sup>(g)</sup> | 8 | 45.3<br>[39.8, 49.4] | 6/8<br>(75%) | 5.6<br>[0.6, 12.5] | 3.0<br>[2.2, 5.0] | 22 / 22 |
| **Total** | 296 | 51.9<br>[39.9, 62.7] | 184/295<br>(62%) | 13.6<br>[6.0, 23.8] | 4.5<br>[3.0, 6.0] | 1644 / 1603 |
| *p*<br>(omnibus) | | $3.5 \times 10^{-11}$ | 0.270 | $1.4 \times 10^{-7}$ | $1.2 \times 10^{-19}$ | |

Values are median [IQR] unless otherwise noted. Superscript letters indicate significant pairwise differences (Holm-corrected Wilcoxon rank-sum test, $p < 0.05$ ): <sup>a</sup>HD, <sup>b</sup>RR-MS, <sup>c</sup>SP-MS, <sup>d</sup>PP-MS, <sup>e</sup>NIND, <sup>f</sup>OIND, <sup>g</sup>CIS/RIS. Omnibus p-values: Kruskal-Wallis (continuous) or chi-squared (sex). Disease duration unavailable for HD. EDSS matched within 7 days of first assessment (available for N = 264/296).

To address the ceiling effects associated with the traditional bedside Luria FEP task, we developed a digital motor sequencing paradigm. Rather than relying on manual hand gestures, this task translates the three-part motor program of the FEP test into sequence-controlled geometric shapes (dots, directional lines, and circles) drawn on a touchscreen interface. This design preserves the essential requirement of dynamic praxis (i.e., sequential transitions between distinct motor programs), while enabling automated, high-resolution performance tracking. By forcing rapid transitions under a timed condition, the paradigm increases the cognitive-motor load, allowing us to decompose dynamic praxis into discrete, measurable subprocesses such as planning latency, trajectory precision, and sequence transition speed.

The digital motor sequencing assessment consists of a 30-second timed trial requiring participants to recall and repeatedly trace a memorized three-symbol sequence on a touchscreen interface (see Methods and Figure 1). This testing paradigm was performed sequentially with both the left and right hands. Each drawing event was classified as correct, incorrect (wrong shape in the sequence), or unrecognized (failed shape detection), and complete sequences were identified via a non-greedy string-matching algorithm (Supplementary Figure S2).

**Figure 1.**
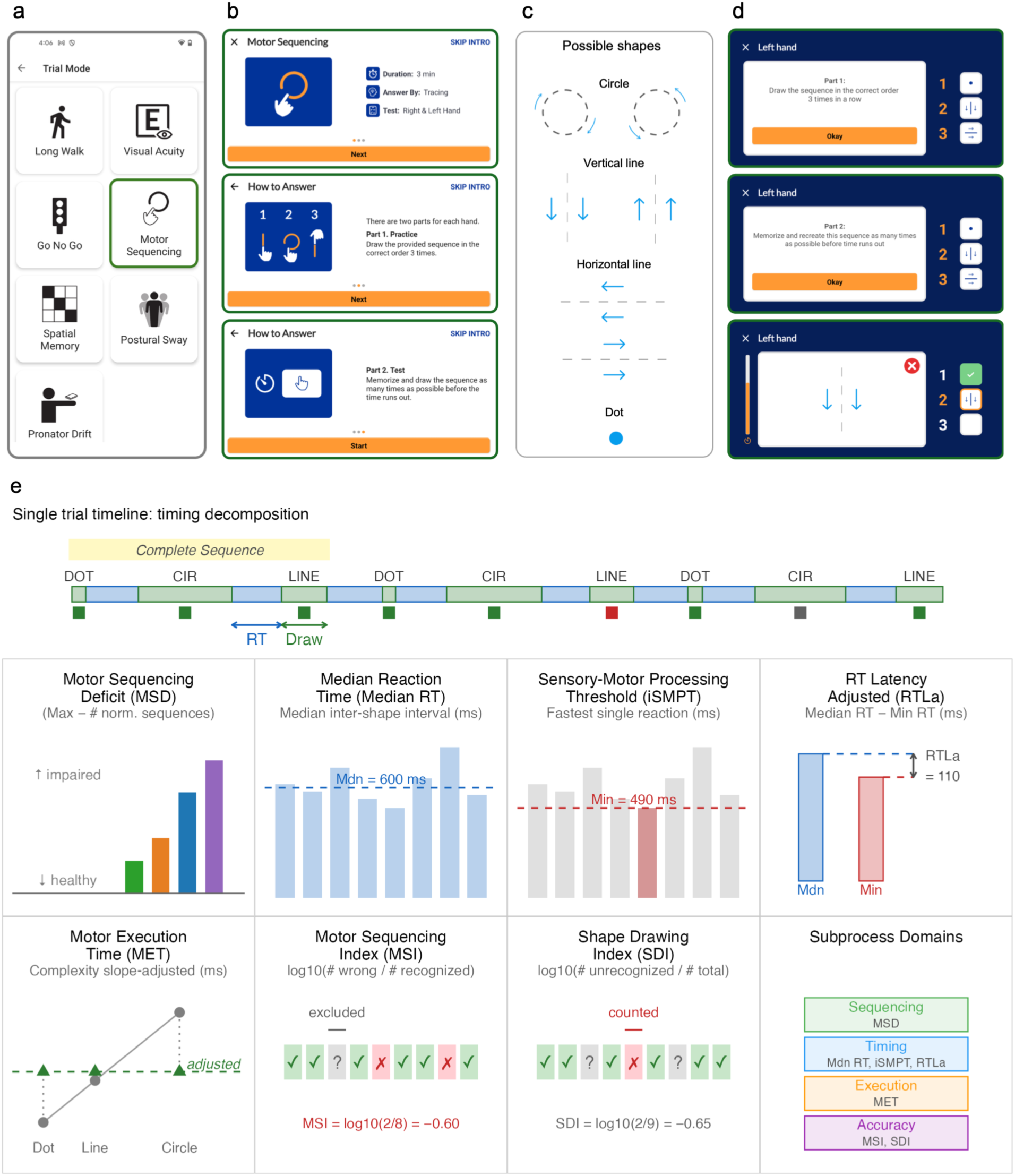
The NeuFun-TS motor sequencing task: app interface, shape vocabulary, trial structure, and biomarker definitions. (a) NeuFun-TS app home screen showing the motor sequencing module alongside other neurological function tests. (b) Introductory instruction screens: task overview (top), guided practice instructions (middle), and timed trial instructions (bottom). (c) The seven directional shapes available for sequence construction: Circle (clockwise/anti-clockwise), Vertical line (downward/upward), Horizontal line (leftward/rightward), and Dot. Blue arrows indicate prescribed drawing direction. (d) Trial execution screenshots (left hand). Top: guided practice with the assigned sequence displayed. Middle: memorization and timed reproduction instructions. Bottom: active trial with real-time feedback (green checkmark = correct; red X = incorrect/unrecognized). (e) Biomarker definitions and subprocess decomposition. Top: timeline of a single trial showing interleaved reaction time (RT; finger-up-to-finger-down) and drawing duration (finger-down-to-finger-up) intervals. A complete sequence (highlighted) is defined as three consecutive correctly drawn shapes representing all three distinct shape types. Middle and bottom: schematic definitions of all seven biomarkers organized by subprocess domain - Sequencing (MSD), Timing (Median RT, iSMPT, RTLa), Execution (MET), and Accuracy (MSI, SDI). NeuFun-TS = Neurological Functional Test Suite; RT = reaction time; MSD = Motor Sequencing Deficit; Median RT = Median Reaction Time; iSMPT = individualized Sensory-Motor Processing Threshold; RTLa = Reaction Time Latency adjusted; MET = Motor Execution Time; MSI = Motor Sequencing Index; SDI = Shape Drawing Index; OLS = ordinary least squares.

Applying the quality control (QC) criteria described in Methods (Supplementary Figure S3a), we excluded 31 HD trials (8.2% of 376; 11 fail-tier plus 20 from one subject-level exclusion). An additional 678 of 3,078 HD+MS trials (22%) were flagged as “suspect” and retained for analysis. A sensitivity analysis applying a more stringent, targeted QC filter confirmed that retaining suspect trials did not inflate group separation metrics (Supplementary Figure S3).

### Timing decomposition separates motor planning from motor execution

Decomposing each inter-shape interval into reaction time (RT; corresponding to pause between drawing shapes and reflecting motor planning and initiation) and drawing duration (reflecting motor execution; Figure 2a) revealed a fundamental dissociation: RT was independent of shape complexity. Median RT was approximately equal across Dot, Line, and Circle targets within each group (HD: 322-332 ms; SP-MS: 640-642 ms), whereas drawing duration showed a strong, monotonic complexity gradient (HD: Dot = 77 ms, Line = 228 ms, Circle = 482 ms, Figure 2b). An event-level analysis across all complete sequences (N = 74,755 events) confirmed this dissociation statistically: RT was nearly invariant to both the previously drawn shape (the non-parametric effect size estimator ε² for a Kruskal-Wallis test ranging from 0 [no variance explained by grouping variable] to 1 [the grouping variable explains all of the variance]; ε^2^ = 0.011, delta median = 56 ms) and the upcoming shape (ε^2^ = 0.004, delta median = 48 ms), whereas drawing duration varied 7-fold across shape types (ε^2^ = 0.716, delta median = 543 ms; Figure 2a,B).

**Figure 2.**
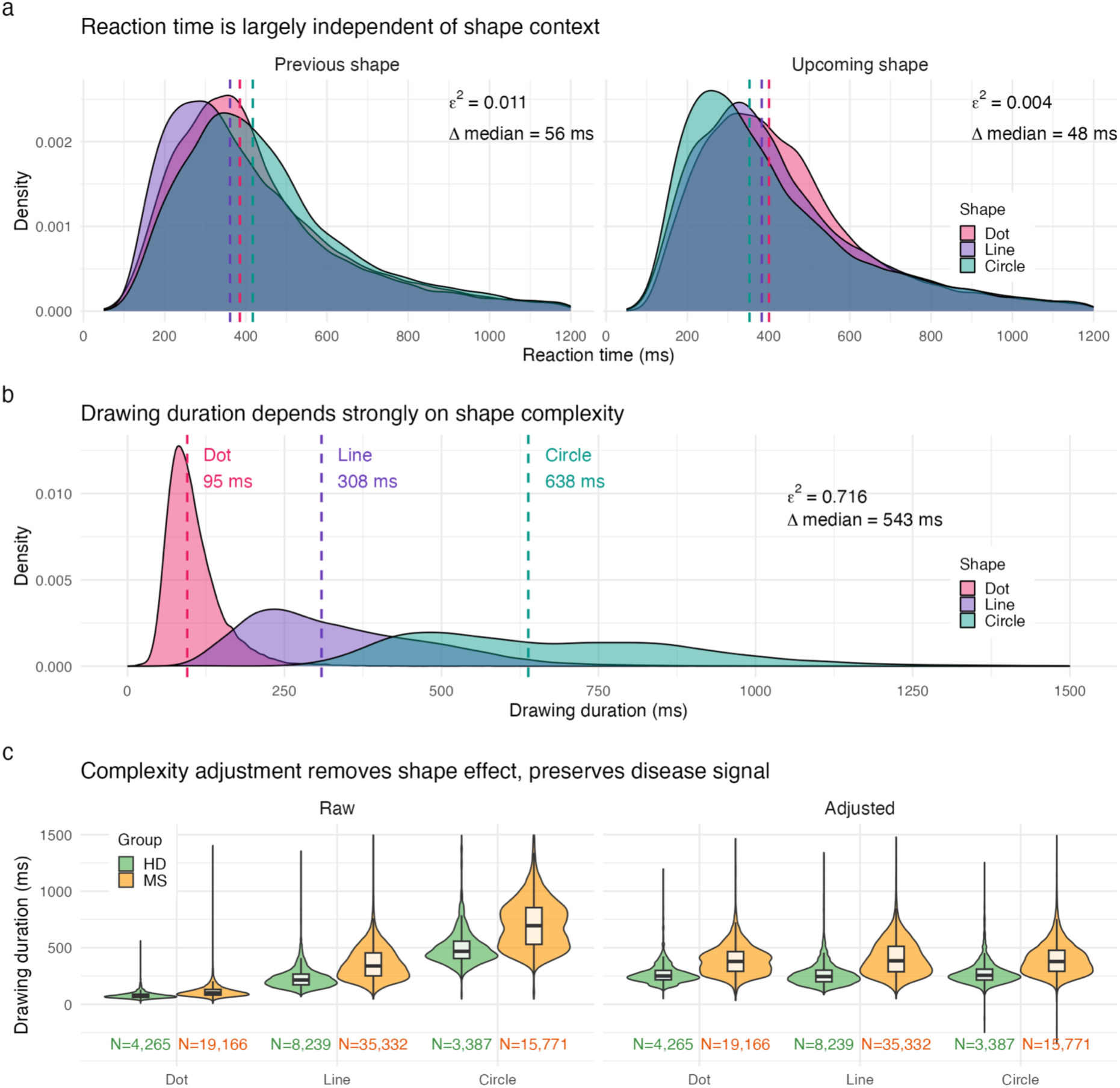
Timing decomposition separates motor planning from motor execution. (a) Density distributions of reaction time stratified by shape context, faceted by previous shape (left) and upcoming shape (right). Reaction time is largely independent of shape context. (b) Density distributions of drawing duration by shape type, pooled across all groups. Drawing duration varies substantially across shape types, demonstrating a strong complexity gradient. (c) Violin plots showing drawing duration by shape type and group (HD, green; MS, orange), faceted by Raw (left) and Adjusted (right). The per-trial OLS complexity adjustment removes shape-dependent variance while preserving the disease signal. HD = healthy donors; MS = multiple sclerosis; RT = reaction time; OLS = ordinary least squares.

The per-trial ordinary least squares (OLS) complexity adjustment (see Methods) removed the shape-dependent variance while preserving the disease signal between HD and MS (Figure 2c). The finding that planning time does not depend on the motor demands of either the preceding or upcoming shape, while execution time does, confirms that our timing decomposition successfully isolates cognitive-motor initiation from motor execution per se.

A difficulty factor correction successfully eliminated composition-dependent variance in sequence counts, enabling pooling across trial types without sacrificing group discrimination. The inter-biomarker correlation matrix confirmed that the seven biomarkers tap into four dissociable motor subprocesses: speed (Median Reaction Time [Median RT], individualized Sensory-Motor Processing Threshold [iSMPT], adjusted Reaction Time Latency [RTLa]), accuracy (Motor Sequencing Index [MSI], Shape Drawing Index [SDI]), motor execution (Motor Execution Time [MET]), and integrative throughput (Motor Sequencing Deficit [MSD]). These validation analyses are detailed in the Supplementary Results (Supplementary Figures S4-S5).

### Six of seven digital biomarkers significantly separate disease groups, with MSD showing the strongest effect

Using first-trial data (one observation per patient-hand, N = 479), we evaluated each biomarker’s ability to separate the four diagnostic groups (Supplementary Table S1; Figure 3a-g). MSD emerged as the strongest discriminator (rank-biserial r = 0.73, Kruskal-Wallis p = 1.1×10^-21^), followed by Median RT (r = 0.69, p = 8.0×10^-21^), RTLa (r = 0.62, p = 3.3×10^-15^), MET (r = 0.50, p = 4.9×10^-11^), iSMPT (r = 0.39, p = 2.0×10^-7^), and MSI (r = 0.36, p = 8.9×10^-5^). SDI (p = 0.14) did not reach significance.

**Figure 3.**
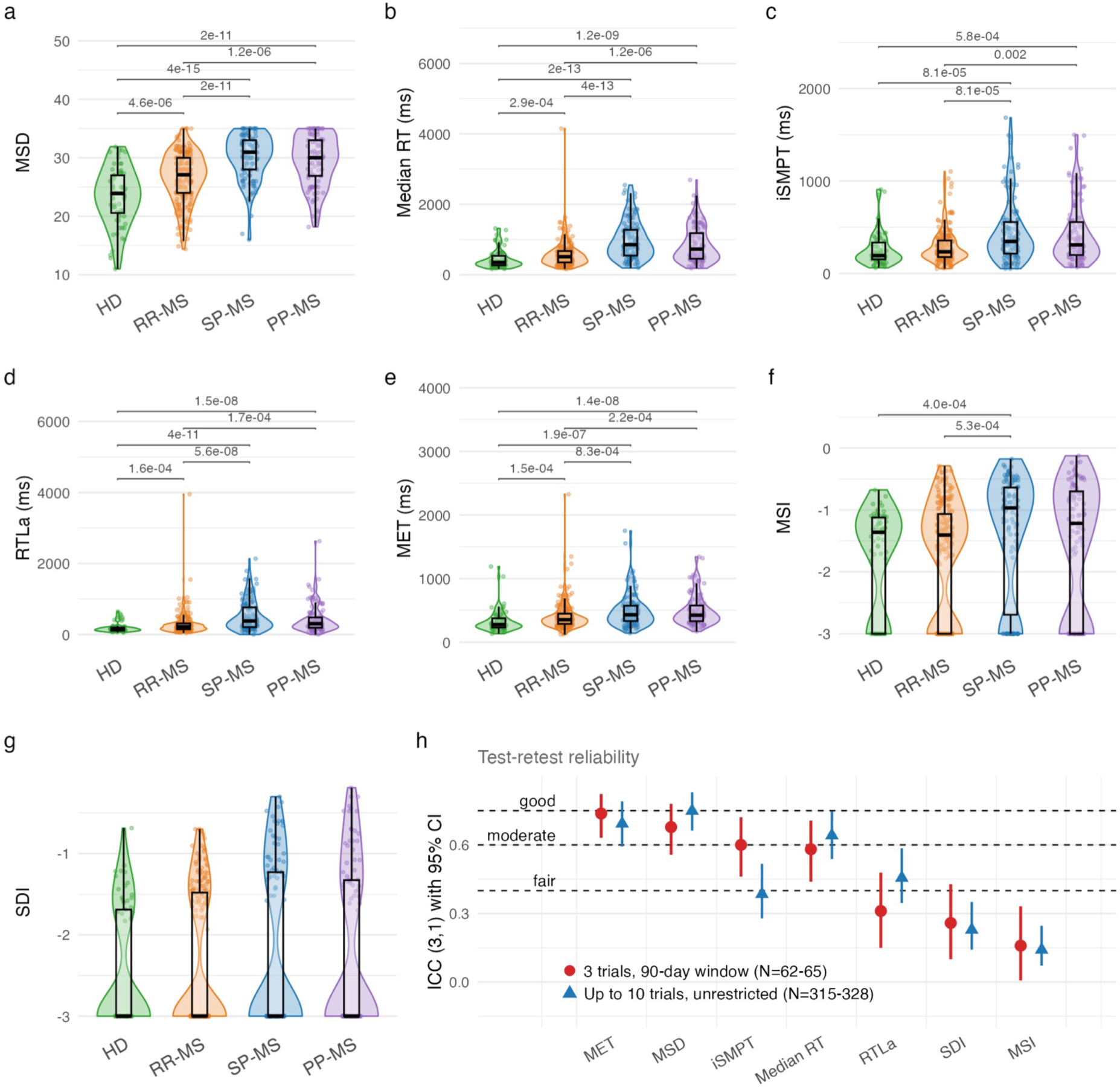
Six of seven biomarkers significantly separate disease groups, with high test-retest reliability for speed and execution measures. (a-g) Violin plots with embedded boxplots showing biomarker distributions across four diagnostic groups (HD, green; RR-MS, orange; SP-MS, blue; PP-MS, purple) using first-trial data. Panels show: (a) MSD, (b) Median RT, (c) iSMPT, (d) RTLa, (e) MET, (f) MSI, (g) SDI. Horizontal brackets indicate significant pairwise comparisons. (h) Forest plot of ICC(3,1) test-retest reliability estimates with 95% confidence intervals for all seven biomarkers under two conditions: first 3 trials within 90 days (red circles) and up to 10 trials unrestricted (blue triangles). Dashed lines indicate conventional ICC thresholds (fair = 0.40, moderate = 0.60, good = 0.75). HD = healthy donors; RR-MS = relapsing-remitting MS; SP-MS = secondary-progressive MS; PP-MS = primary-progressive MS; MSD = Motor Sequencing Deficit; Median RT = Median Reaction Time; iSMPT = individualized Sensory-Motor Processing Threshold; RTLa = Reaction Time Latency adjusted; MET = Motor Execution Time; MSI = Motor Sequencing Index; SDI = Shape Drawing Index; ICC = intraclass correlation coefficient.

Across all significant biomarkers, median values demonstrated a consistent ordinal gradient: HD performed best, followed by RR-MS, with SP-MS and PP-MS showing the greatest impairment. For example, Median RT was 351 ms in HD versus 854 ms in SP-MS (i.e., a 2.4-fold slowing) while MSD was 23.9 in HD versus 30.9 in SP-MS (a 29% increase in motor sequencing deficit). Pairwise Wilcoxon comparisons revealed that HD separated from all MS groups, RR-MS separated from progressive MS, and SP-MS and PP-MS were generally indistinguishable from each other.

To evaluate whether biomarker sensitivity was specific to MS-related pathology or reflected generic neurological illness, we extended the analysis to include participants with non-inflammatory neurological disease (NIND, N = 50), other inflammatory neurological disease (OIND, N = 42), and clinically/radiologically isolated syndrome (CIS/RIS, N = 16; Supplementary Figure S6). HD and NIND showed comparable performance and separated clearly from MS, OIND partially overlapped with MS groups, and CIS/RIS occupied an intermediate position between HD and established MS.

### MET and MSD achieve the highest test-retest reliability; accuracy measures are limited by floor effects

To assess measurement stability, we computed the intraclass correlation coefficient (ICC) from the first three trials per patient-hand completed within a 90-day window (N = 62-65 subjects; Supplementary Table S2; Figure 3h). MET achieved the highest reliability (ICC = 0.74 [95% confidence interval (CI): 0.63, 0.82]), followed by MSD (ICC = 0.68 [0.56, 0.78]), both placing in the “moderate” range. iSMPT (ICC = 0.60 [0.46, 0.72]) reached the “moderate” threshold, while Median RT (ICC = 0.58 [0.44, 0.71]) fell in the “fair” range. RTLa (ICC = 0.31 [0.15, 0.48]), SDI (ICC = 0.26 [0.10, 0.43]), and MSI (ICC = 0.16 [0.01, 0.33]) were “poor.”

The reliability hierarchy reflects the nature of each biomarker: motor execution speed (MET) and integrative throughput (MSD) are inherently stable because they aggregate information across many events within a trial, whereas accuracy measures depend on rare error events. The poor ICC of MSI likely reflects a floor effect, as many trials have zero sequence errors, limiting between-subject variance relative to within-subject fluctuation. MSD, as a deficit from a theoretical maximum, is a linear transformation of raw throughput that preserves the original variance structure while providing an intuitive scale where 0 represents theoretically perfect performance and 35 represents zero throughput.

Consistent with this moderate reliability, averaging across repeat trials improved group separation (Supplementary Table S3): comparing effect sizes from the single first trial (N = 479) versus the mean of the first three trials within 90 days (N = 216) showed that averaging improved discrimination in five of seven biomarkers, with the largest gain for SDI (+0.46). This confirms that repeat testing reduces measurement noise and supports recommending multi-trial assessment when clinically feasible.

### The Motor Sequencing Composite achieves high concordance index for diagnostic classification

We asked whether combining motor sequencing biomarkers into a single composite score (i.e., the Motor Sequencing Composite [MSC]) could outperform any individual metric in an ordinal classification of diagnoses based on the level of brain dysfunction: HD < RR-MS < progressive MS (P-MS). The training set comprised 358 observations (180 patients) and the held-out validation set 185 observations (94 patients), stratified by group, age, sex, and NeurEx (Neurological Examination) total (Figure 4a).

**Figure 4.**
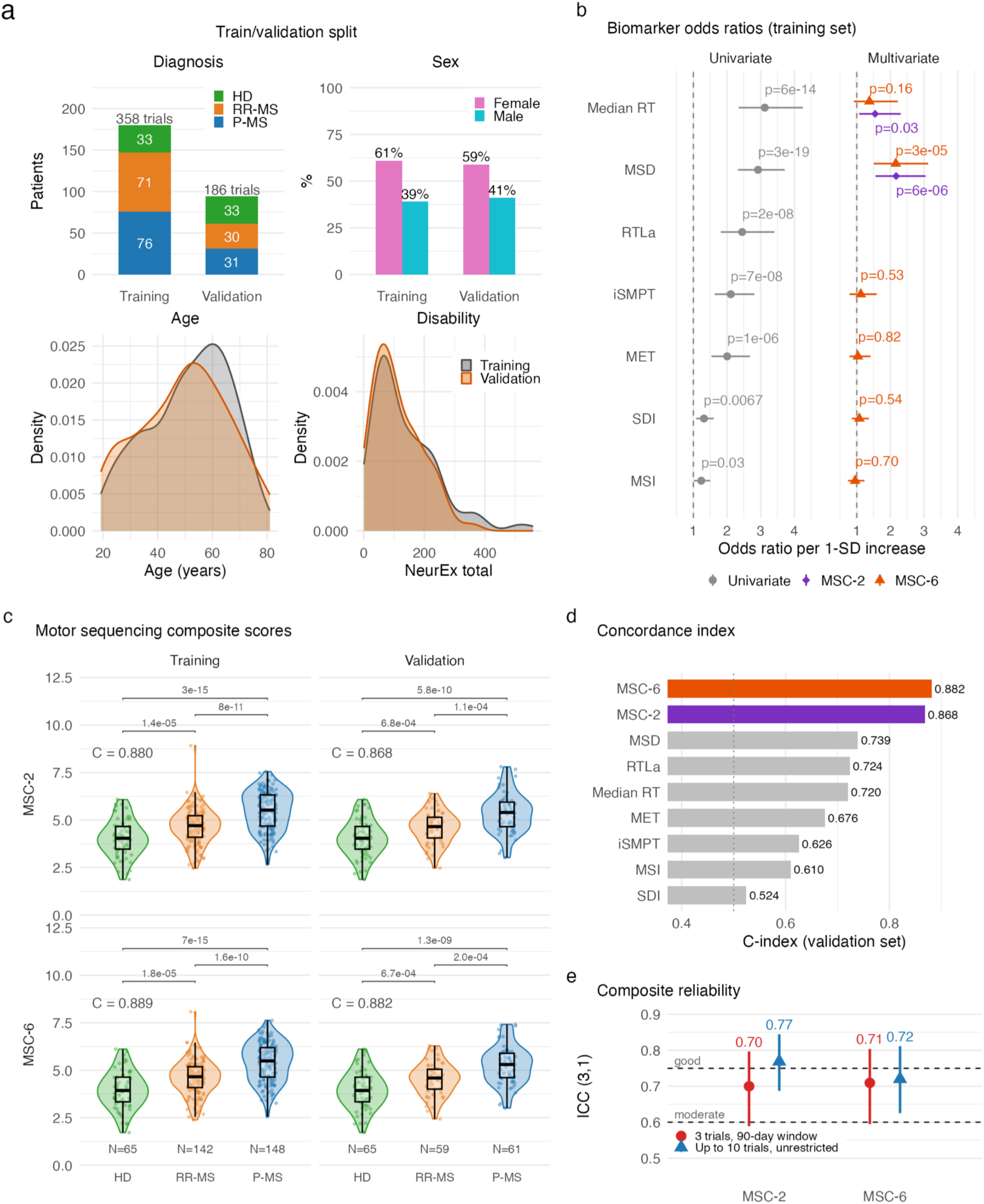
Motor Sequencing Composites (MSC-2 and MSC-6) achieve excellent concordance for MS severity classification. (a) Train/validation split overview showing balanced stratification by group, sex, age, and NeurEx total across Training and Validation sets. (b) Forest plot of biomarker odds ratios per 1-SD increase. Left: univariate odds ratios. Right: multivariate odds ratios from MSC-6 (full model) and MSC-2 (stepwise model). (c) Violin plots showing OLR linear predictor by diagnostic group for MSC-2 (top) and MSC-6 (bottom) across Training (left) and Validation (right) sets, with C-index values annotated. (d) Concordance index comparison on the validation set for individual biomarkers versus MSC-2 and MSC-6. (e) Test-retest reliability (ICC with 95% CI) of MSC-2 and MSC-6 under two conditions: 3 trials within 90 days (red circles) and up to 10 trials unrestricted (blue triangles). Dashed lines indicate moderate (0.60) and good (0.75) thresholds. MSC-2 = Motor Sequencing Composite (2-predictor: MSD, Median RT); MSC-6 = Motor Sequencing Composite (6-predictor); HD = healthy donors; RR-MS = relapsing-remitting MS; P-MS = progressive MS; OLR = ordinal logistic regression; C-index = concordance index; ICC = intraclass correlation coefficient; MSD = Motor Sequencing Deficit; Median RT = Median Reaction Time; NeurEx = Neurological Examination.

RTLa was excluded from the candidate set due to strong collinearity with Median RT and iSMPT, yielding the full six-predictor model (MSC-6: MSD, Median RT, iSMPT, MET, MSI, SDI). Forward stepwise Akaike Information Criterion (AIC) selection from MSC-6 identified a parsimonious two-variable model (MSC-2: MSD, Median RT; Figure 4b).

In the independent validation set (Supplementary Table S4; Figure 4c), both MSC-2 and MSC-6 achieved excellent discriminatory capacity. MSC-2 achieved a concordance index of 0.868 (C-index ranging from 0-1, which generalizes the Area Under the Receiver Operator Curve [AUROC] to quantify the model’s ability to discriminate between ordered diagnostic categories), while MSC-6 yielded a C-index of 0.882 (Figure 4d). Both composites most frequently misclassified RR-MS as P-MS. This directional error aligns with the clinical reality that discrete categorical stratification fails to capture the underlying biological complexity of continuous, overlapping phenotypic processes, such as accumulation of physical and cognitive disabilities.

Both composites demonstrated good test-retest reliability (Figure 4e): MSC-2 achieved ICC = 0.70 [95% CI: 0.58, 0.80] and MSC-6 achieved ICC = 0.71 [0.60, 0.80] in the primary analysis (first 3 trials within 90 days, N = 65), improving to ICC = 0.77 [0.69, 0.84] and 0.72 [0.63, 0.81] respectively in the secondary analysis (up to 10 trials, unrestricted time span, N = 328). These values exceed the individual biomarker ICCs, demonstrating that combining complementary measures stabilizes the measurement.

### Complexity × group interaction reveals a capacity-limited motor execution buffer

Our shape-complexity slope adjustment normalizes drawing duration to remove the effect of shape type. However, we reasoned that the slope itself (i.e., the additional milliseconds imposed by each unit increase in complexity) might be an informative biomarker: if MS patients have a capacity-limited motor programming buffer, they should pay a disproportionate cost for complex shapes, producing steeper slopes than healthy donors.

This prediction was confirmed as the strongest finding among our six Luria-derived analyses (Figure 5a,b; Supplementary Table S5). The complexity slope was 448 ms/level in HD, 570 in RR-MS, 685 in SP-MS, and 700 in PP-MS (Kruskal-Wallis H = 227.2, p = 5.6×10^-49^). Pairwise contrasts separated all MS groups from HD, RR-MS from both progressive groups, while SP-MS and PP-MS did not differ from each other.

**Figure 5.**
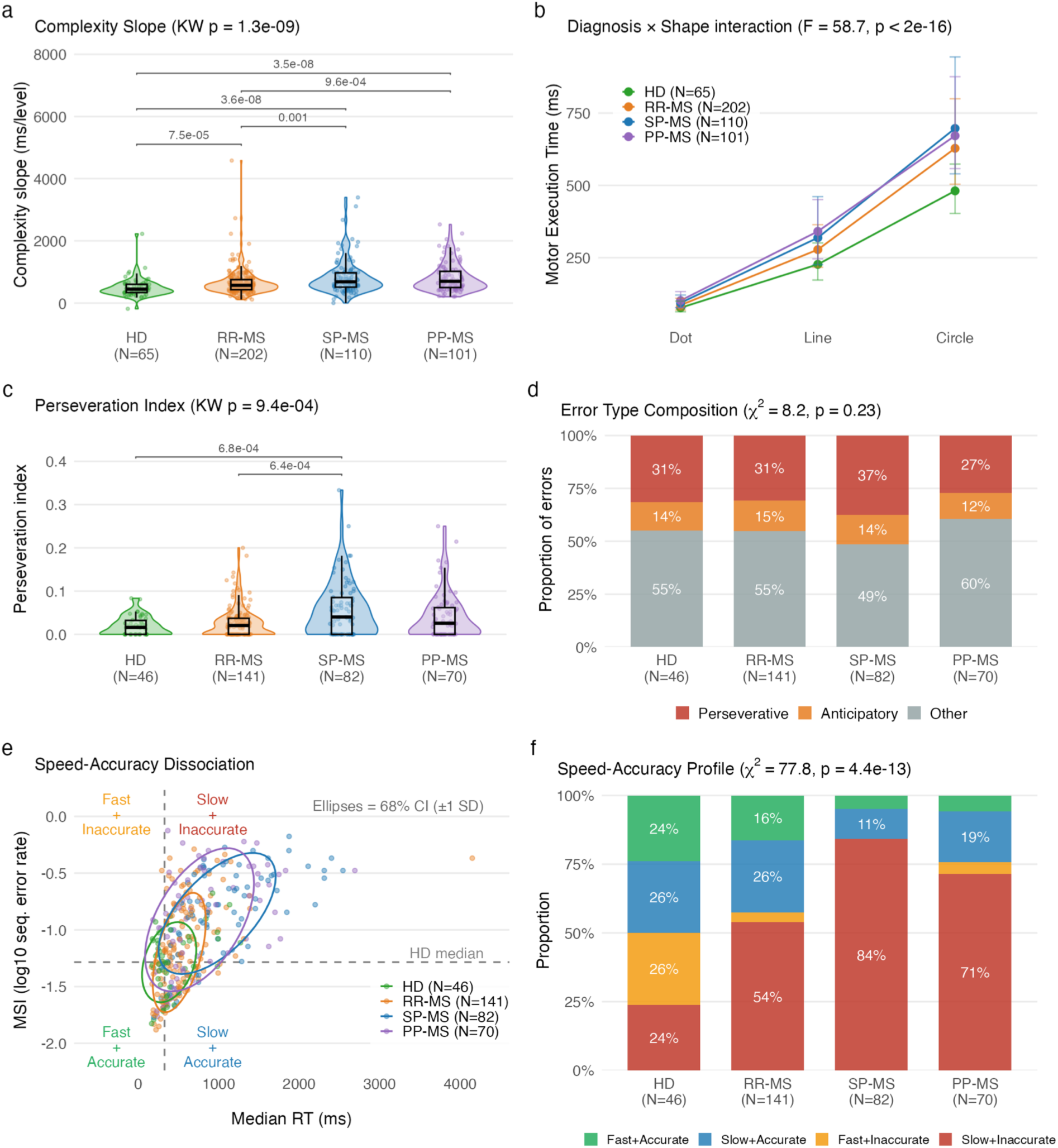
Luria-derived analyses reveal motor capacity limitation, perseveration, and speed-accuracy dissociation. (a) Violin plots showing complexity slope (ms/level) by diagnostic group. All MS groups show steeper slopes than HD, indicating disproportionate motor cost for complex shapes. (b) Median Motor Execution Time by shape complexity (Dot, Line, Circle) for each group with IQR error bars, demonstrating a significant Diagnosis × Shape interaction. (c) Violin plots of Perseveration Index by group, restricted to patient-hands with at least one error. SP-MS shows elevated perseverative error frequency. (d) Stacked bar chart of error type composition (Perseverative, Anticipatory, Other) by group. The proportion of perseverative errors is stable across groups, indicating MS increases error frequency without changing the qualitative error profile. (e) Speed-accuracy dissociation scatter plot (Median RT vs MSI) with 68% confidence ellipses per group. Quadrants defined relative to HD medians. Progressive MS groups cluster in the Slow+Inaccurate quadrant. (f) Stacked bar chart of speed-accuracy quadrant proportions by group. SP-MS is predominantly Slow+Inaccurate, while PP-MS shows a higher proportion of Slow+Accurate (compensating) pattern. HD = healthy donors; RR-MS = relapsing-remitting MS; SP-MS = secondary-progressive MS; PP-MS = primary-progressive MS; MET = Motor Execution Time; RT = Reaction Time; MSI = Motor Sequencing Index; IQR = interquartile range.

Crucially, this interaction was motor execution-specific: RT showed no shape × group interaction, implying that all shapes required approximately equal planning time within each group. This dissociation demonstrates that MS does not simply cause generalized motor slowing but specifically impairs the capacity to execute complex motor programs. The finding is analogous to capacity-limitation effects in the Luria literature, where more complex sequences (alternating patterns) produce disproportionate difficulty for frontal-subcortical patients compared to simple repetitive movements.

### Perseveration index validates the “digital Luria” construct

The hallmark of frontal-subcortical dysfunction on the Luria FEP test is not simply making errors but making perseverative errors - i.e., repeating a previously executed motor program instead of advancing to the next sequence step. To test whether our smartphone task captures this specific deficit, we classified each sequence error as perseverative (drew previous shape), anticipatory (drew next shape), or other.

The proportion of errors that were perseverative was remarkably stable across all groups (33-36%), indicating that MS does not change the qualitative type of error. However, restricting to patient-hands with at least one error (N = 339; see Methods), the absolute frequency of perseverative errors per trial differed significantly across groups (Kruskal-Wallis H = 10.9, p = 0.013; Figure 5c,d): SP-MS showed mean perseveration index = 0.058 versus HD = 0.020, with the effect driven by SP-MS being elevated relative to both HD (p = 0.0007) and RR-MS (p = 0.0006). PP-MS did not differ from HD or RR-MS.

### Speed-accuracy dissociation reveals distinct deficit subtypes across MS phenotypes

We hypothesized that the digitalized motor sequencing task can decompose performance into distinct subprocesses, enabling identification of where in the cognitive-motor chain performance breaks down. We tested this directly by plotting each patient in a two-dimensional speed × accuracy space (median RT vs MSI; quadrants defined relative to HD medians; Figure 5e). We restricted this analysis to patient-hands with at least one sequencing error (N = 339; see Methods), because zero-error patients (MSI = −3 floor) have perfect accuracy by definition and do not participate in the speed-accuracy tradeoff.

In HD, speed and accuracy were weakly and non-significantly correlated (Spearman rho = 0.23, p = 0.12). In MS, the correlation became strongly positive: rho = 0.55 in RR-MS (p = 1.0×10^-12^) and rho = 0.65 in SP-MS (p = 4.4×10^-11^). This coupling of speed and accuracy (i.e., slower patients are also making more errors) suggests a capacity limitation in which subject attempts but cannot compensate for the underlying deficit by prolonging motoric planning. The quadrant distribution confirmed this pattern (χ^2^ = 77.8, df = 9, p = 4.4×10^-13^): 84.1% of SP-MS fell in the “Slow + Inaccurate” quadrant versus 23.9% of HD (Figure 5f).

PP-MS showed a notably different pattern from SP-MS despite similar overall disability: a higher proportion occupied the “Slow + Accurate” (compensating) quadrant (18.6% vs 11.0%). We interpret this as reflecting the more focal pyramidal-tract involvement in PP-MS slowing motor execution while preserving cognitive motor programming, versus the more diffuse cerebral pathology in SP-MS that compromises both speed and sequence accuracy. Semi-quantitative magnetic resonance imaging (MRI) data corroborated this interpretation (see Figure 6b).

**Figure 6.**
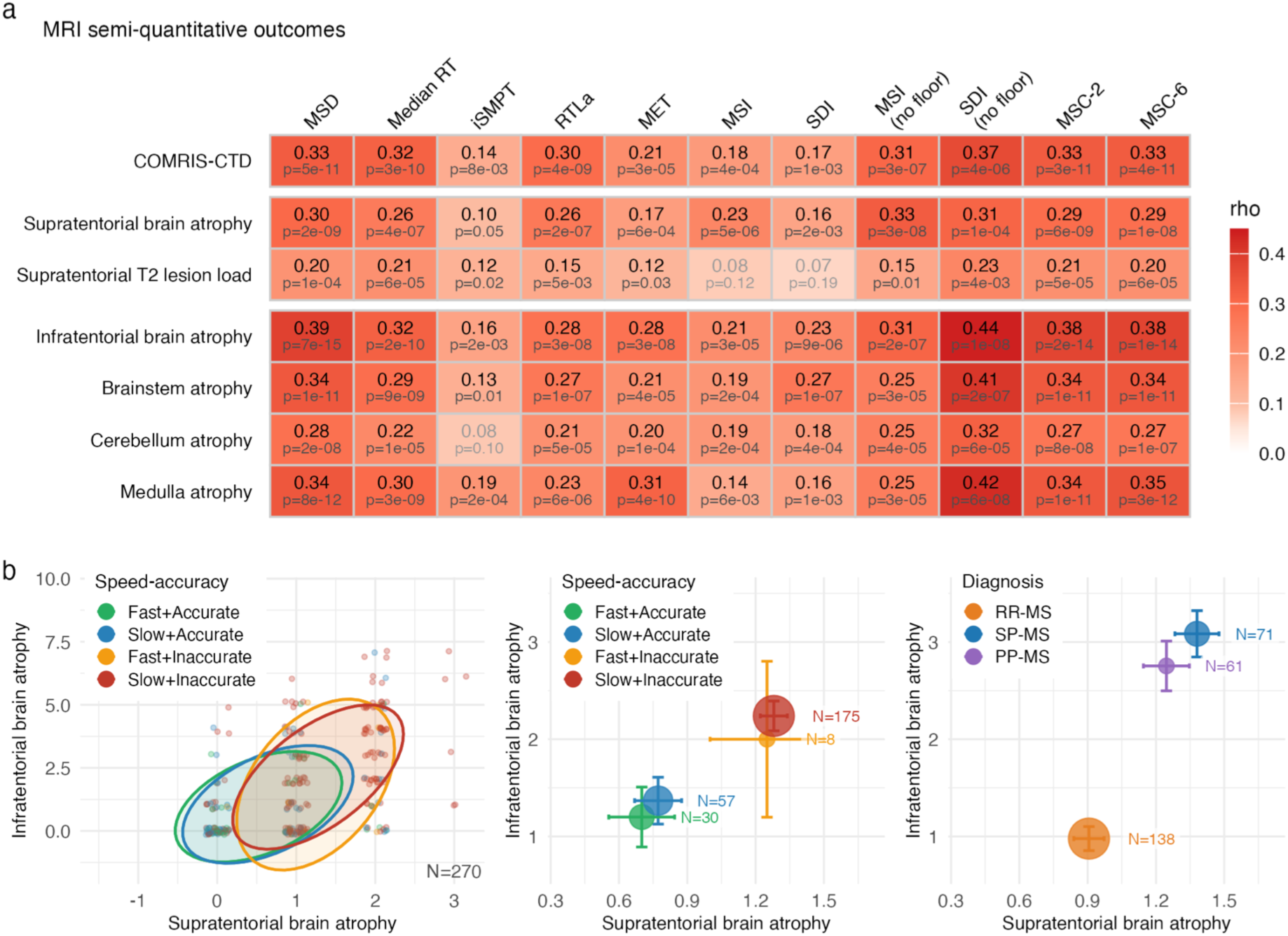
Clinical validation: MRI structural outcomes and regional atrophy patterns. (a) Heatmap of Spearman rank correlations between motor sequencing biomarkers (columns) and semi-quantitative MRI outcomes (rows) in MS patients. MRI outcomes are organized into three blocks: global disability (COMRIS-CTD), supratentorial measures (brain atrophy, T2 lesion load), and infratentorial atrophy measures (composite, brainstem, cerebellum, medulla). Each cell shows Spearman rho and p-value. (b) Supratentorial brain atrophy versus infratentorial brain atrophy composite in three views. Left: scatter plot with 68% confidence ellipses colored by speed-accuracy quadrant. Middle: group centroids (mean ± SE) by speed-accuracy quadrant. Right: group centroids by MS diagnosis, showing that progressive MS subtypes have higher atrophy burden than RR-MS across both compartments. MRI = magnetic resonance imaging; COMRIS-CTD = Combinatorial MRI Scale of CNS Tissue Destruction; RR-MS = relapsing-remitting MS; SP-MS = secondary-progressive MS; PP-MS = primary-progressive MS; MSD = Motor Sequencing Deficit; MSC-2 = Motor Sequencing Composite (2-predictor: MSD, Median RT); MSC-6 = Motor Sequencing Composite (6-predictor); SE = standard error.

### Informative null results support task design validity

Three additional Luria-derived analyses produced non-significant results that inform task interpretation. First, switch cost: the difference in RT between shape-transition and shape-repeat events did not differ across groups (p = 0.57), indicating that MS slowing is generalized across all transitions rather than specific to the “switching” component of motor programming. This refocuses interpretation on motor execution capacity rather than cognitive set-shifting.

Second, within-trial deceleration was not significant (p = 0.81): all groups predominantly accelerated during the 30-second trial (warm-up effect), confirming that the trial duration does not produce micro-fatigue. This finding directly supports the task’s suitability for repeated unsupervised monitoring - a 30-second trial is brief enough that performance does not degrade even in progressive MS.

Third, error clustering did not differ across groups (p = 0.69), with errors manifesting as isolated events (mean run length ∼1.2-1.3) rather than cascading failures. This lack of compounding errors points to a fundamental characteristic of the MS cognitive phenotype, which diverges sharply from classic dementing illnesses. While cortical and frontostriatal dementias are marked by executive collapse with prominent motor or cognitive perseverations, cognitive impairment in MS is dominated by slowed processing speed with relative preservation of error monitoring and rule maintenance (Rao et al., 1991).

### Motor sequencing biomarkers correlate with MRI measures of structural brain damage

We next evaluated whether the digital biomarkers reflected underlying structural pathology by correlating them with seven semi-quantitative MRI outcomes in MS patients (N = 378; Figure 6a). Among the three tiers of MRI outcomes, infratentorial atrophy measures showed the strongest and most consistent associations with motor sequencing biomarkers, with SDI (no floor) reaching rho = 0.44 for the infratentorial brain atrophy composite. Combinatorial MRI Scale of central nervous system (CNS) Tissue Destruction (COMRIS-CTD), a global measure of CNS tissue destruction, showed moderate correlations across all biomarkers: MSD at rho = 0.33 (p = 5×10^-11^), Median RT at rho = 0.32 (p = 3×10^-10^), and RTLa at rho = 0.30 (p = 4×10^-9^). Supratentorial brain atrophy showed similar associations (MSD: rho = 0.30, Median RT: rho = 0.26). The MSC-2 and MSC-6 composites showed uniformly strong associations across all MRI outcomes.

To explore whether the speed-accuracy dissociation maps onto distinct patterns of regional brain damage, we plotted supratentorial brain atrophy against the infratentorial brain atrophy composite (Figure 6b; N = 270, excluding patients with MSI at the floor value of -3). Patients classified as Slow+Inaccurate showed the highest burden across both regional dimensions, with progressive separation along both axes from Fast+Accurate through compensating and impulsive profiles to the global deficit quadrant. When grouped by diagnosis, progressive MS subtypes showed higher atrophy burden than RR-MS across both supratentorial and infratentorial compartments.

### Neurological examination correlations confirm construct validity

Correlations between digital biomarkers and clinician-derived disability scores plus five selected NeurEx panels (N = 392, matched within 7 days; Figure 7a) revealed the expected pattern. Among clinician-derived scores (Expanded Disability Status Scale [EDSS], Scripps Neurological Rating Scale [SNRS], Combinatorial Weight-adjusted Disability Scale [CombiWISE], NeurEx total), the MSC-2 and MSC-6 composites achieved the strongest correlations (rho = 0.44-0.52 across all clinician-derived measures). Among NeurEx subscores, the strongest associations emerged for cognitive function (MSD: rho = 0.47; MSC-2: rho = 0.48), cerebellar functions (Median RT: rho = 0.47; MSC-2: rho = 0.47), and pyramidal/motor function (Median RT: rho = 0.41; MSC-2: rho = 0.42). Upper extremity strength and pupils/vision panels showed moderate correlations with speed biomarkers. MSC-2 and MSC-6 achieved the highest correlations across all panels, confirming the added value of the composite approach.

**Figure 7.**
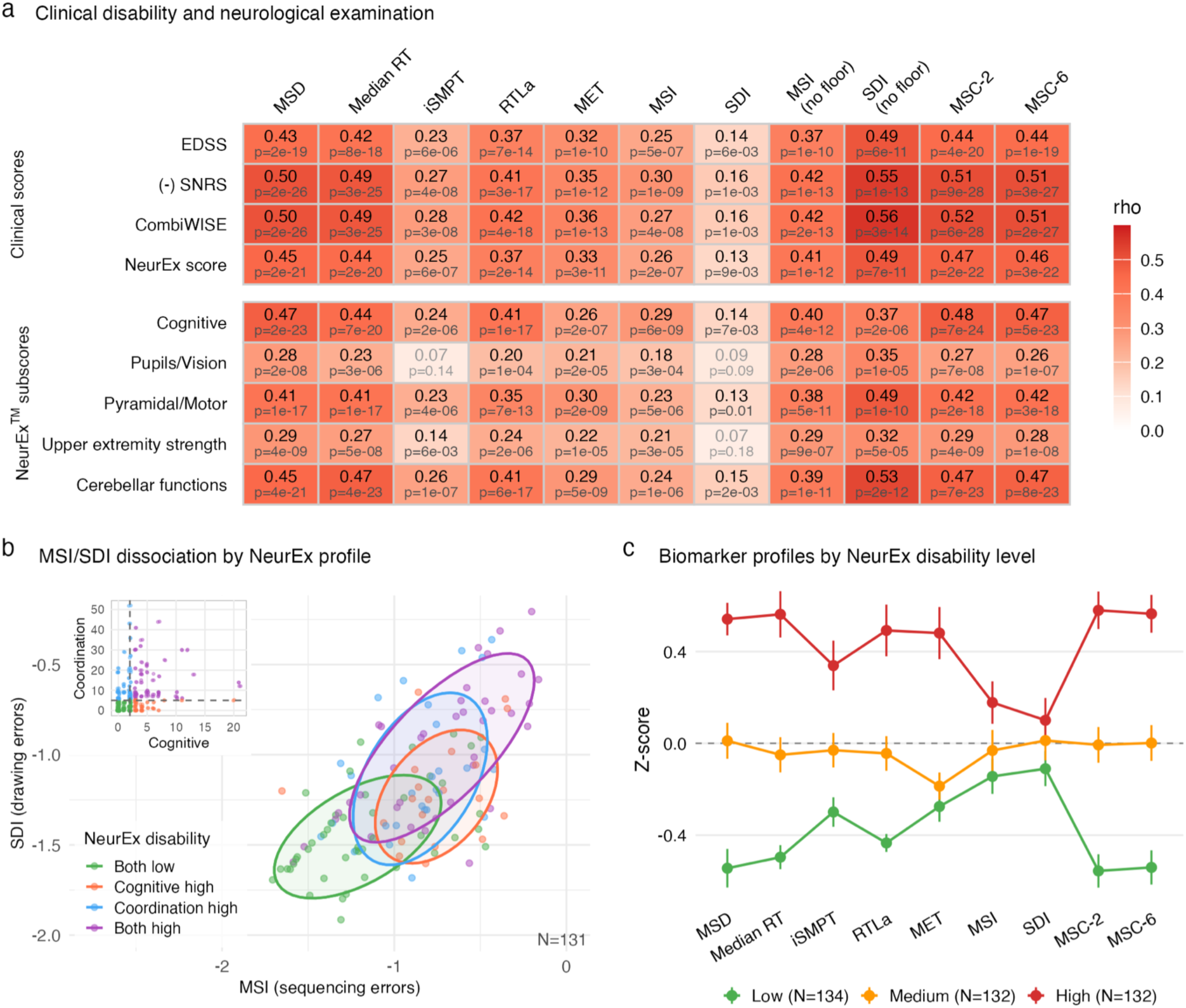
Clinical validation: neurological examination correlations and biomarker dissociation by disability profile. (a) Heatmap of Spearman rank correlations between motor sequencing biomarkers (columns) and clinical/neurological measures (rows) in MS patients matched within 7 days. Rows are divided into clinician-derived scores (EDSS, negated SNRS, CombiWISE, NeurEx score) and NeurEx subscores (Cognitive, Pupils/Vision, Pyramidal/Motor, Upper extremity strength, Cerebellar functions). Each cell shows Spearman rho and p-value. (b) MSI versus SDI scatter plot for patients with both scores above floor, colored by NeurEx disability quadrant (defined by median splits of Cognitive and Coordination subscores). Patients with high coordination disability cluster toward worse SDI, while those with high cognitive disability cluster toward worse MSI. Inset: NeurEx Cognitive versus Coordination scores with quadrant boundaries. (c) Mean Z-scored biomarker profiles across NeurEx total score tertiles (Low, Medium, High), showing a monotonic severity gradient across all biomarkers. MS = multiple sclerosis; EDSS = Expanded Disability Status Scale; SNRS = Scripps Neurological Rating Scale; CombiWISE = Combinatorial Weight-adjusted Disability Scale; NeurEx = Neurological Examination; MSI = Motor Sequencing Index; SDI = Shape Drawing Index; MSC-2 = Motor Sequencing Composite (2-predictor: MSD, Median RT); MSC-6 = Motor Sequencing Composite (6-predictor); SE = standard error.

The difference in correlations of NeurEx subdomains with MSI and SDI further supported construct validity of the motor sequencing performance decomposition. To directly test whether sequencing errors and drawing errors reflect distinct neurological substrates, we examined patients with both MSI and SDI above floor (N = 131; Figure 7b), stratified by NeurEx disability profile into four quadrants defined by median splits of cognitive (Cognitive score > 2) and coordination (Coordination score > 5) subscores. Patients with high coordination disability clustered toward worse SDI (drawing errors), while patients with high cognitive disability clustered toward worse MSI (sequencing errors), demonstrating domain-specific dissociation between the two accuracy biomarkers.

To test whether digital biomarker performance tracks neurological disability burden in a dose-response manner, we stratified MS patients into tertiles of total NeurEx score (Low, Medium, High) and computed z-scored biomarker profiles for each group (Figure 7c). All biomarkers showed a monotonic gradient across severity tertiles, with the High-severity group performing approximately 0.4-0.8 standard deviations worse than the Low-severity group. MSD, Median RT, MSC-2, and MSC-6 showed the steepest severity gradients, while accuracy measures (MSI, SDI) showed flatter profiles - consistent with their floor effects limiting dynamic range. This dose-response relationship confirms that the digital biomarkers capture a continuous gradient of neurological impairment rather than merely distinguishing diagnostic categories.

### Cognitive measures confirm convergent validity across independently derived tasks

Correlations with Neurological Functional Test Suite (NeuFun-TS) cognitive measures (N = 293 complete cases; Figure 8a) demonstrated broad convergent validity. The Symbol Digit Modalities Test (SDMT) showed the strongest correlations (MSD: rho = 0.59, Median RT: rho = 0.58, MSC-2: rho = 0.61, MSC-6: rho = 0.61), consistent with both tasks requiring rapid information processing under time pressure. Among spatial memory latency biomarkers derived from the validated NeuFun-TS spatial memory test (Kosa et al., 2026), iSMPT-S measures showed the highest correlations (Static iSMPT-S vs SDI no floor: rho = 0.50; Dynamic iSMPT-S vs SDI no floor: rho = 0.39), consistent with shared sensory-motor processing demands. The spatial memory composite and predicted SDMT achieved rho = 0.49-0.55 with MSC-2/MSC-6, demonstrating within-platform cross-task convergent validity while confirming that motor sequencing captures additional variance beyond what memory tasks provide.

**Figure 8.**
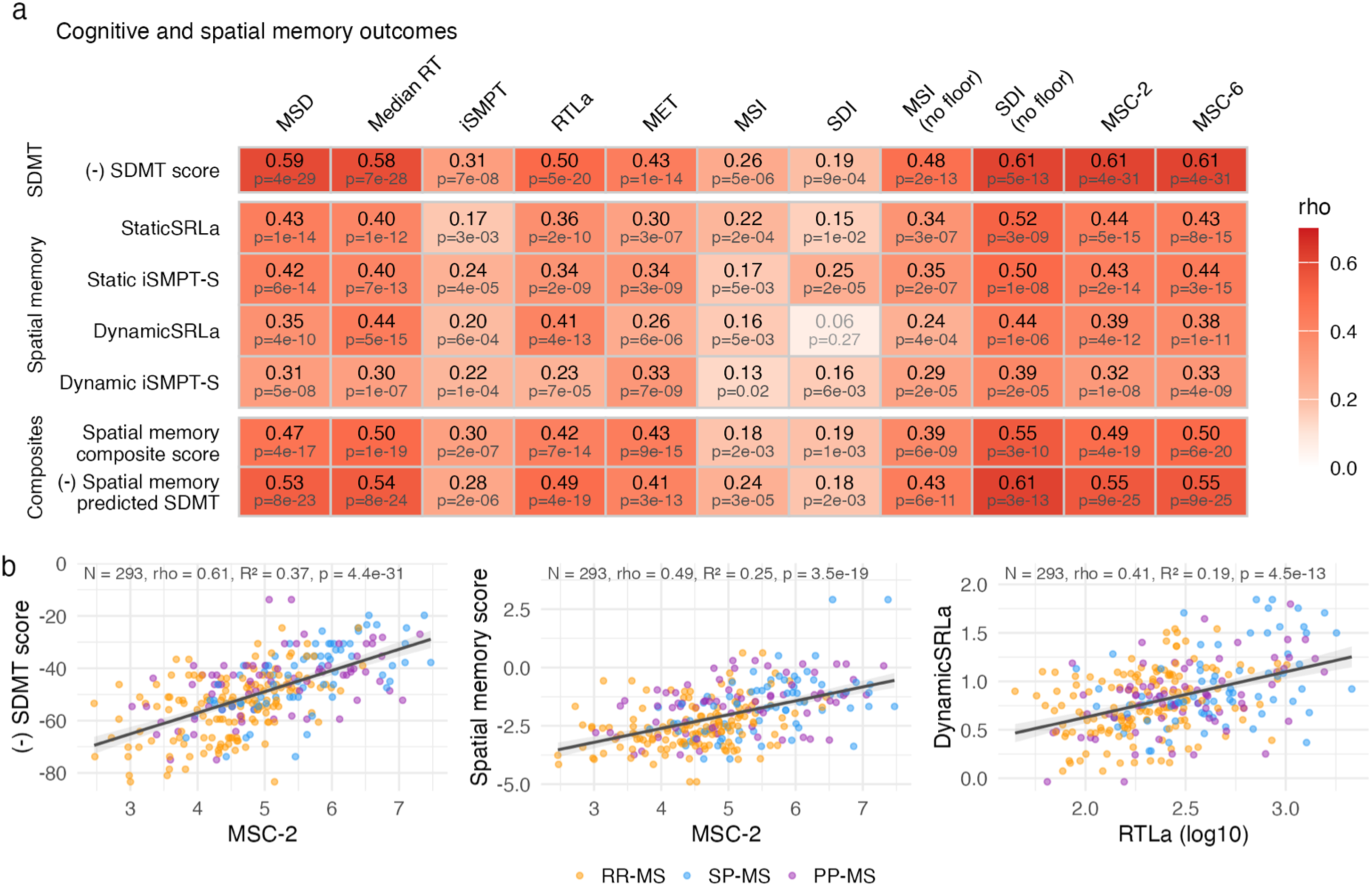
Clinical validation: cognitive and spatial memory convergent validity. (a) Heatmap of Spearman rank correlations between motor sequencing biomarkers (columns) and cognitive/spatial memory outcomes (rows) in MS patients. Outcomes are organized into three sections: SDMT (negated score from NeuFun-TS), Spatial memory latency biomarkers (StaticSRLa, Static iSMPT-S, DynamicSRLa, Dynamic iSMPT-S), and Composites (spatial memory composite, predicted SDMT). Each cell shows Spearman rho and p-value. (b) Three scatter plots with linear regression lines and 95% confidence bands, colored by MS subtype: MSC-2 vs negated SDMT, MSC-2 vs spatial memory composite, and RTLa (log10) vs DynamicSRLa. The first two demonstrate cross-task convergent validity at the composite level; the third demonstrates subprocess-level convergence between analogous cognitive planning latency measures from independently derived tasks. MS = multiple sclerosis; RR-MS = relapsing-remitting MS; SP-MS = secondary-progressive MS; PP-MS = primary-progressive MS; SDMT = Symbol Digit Modalities Test; NeuFun-TS = Neurological Functional Test Suite; MSC-2 = Motor Sequencing Composite (2-predictor: MSD, Median RT); MSC-6 = Motor Sequencing Composite (6-predictor); RTLa = Reaction Time Latency adjusted; StaticSRLa = Static Spatial Reaction Latency adjusted; DynamicSRLa = Dynamic Spatial Reaction Latency adjusted; iSMPT-S = individualized Sensory-Motor Processing Threshold (Spatial memory task).

Three scatter plots (Figure 8b) further illustrated convergent validity at the composite and subprocess levels: MSC-2 correlated with negated SDMT score (rho = 0.61, R^2^ = 0.37, p = 4.4×10^-31^) and with the spatial memory composite (rho = 0.49, R^2^ = 0.25, p = 3.5×10^-19^). At the subprocess level, RTLa (log10-transformed cognitive-motor planning latency) correlated with its spatial memory analogue DynamicSRLa (rho = 0.41, R^2^ = 0.19, p = 4.5×10^-13^), demonstrating that analogous cognitive subprocesses extracted from independently designed tasks converge on a shared underlying construct.

## Discussion

Throughout adult human life, neurological functions are largely unmeasured, creating a clinical blind spot that delays disease detection until pathological decline is well underway. Standard neurological evaluation typically brackets the lifespan, focusing on developmental milestone acquisition in pediatrics and reactive diagnostic workups after symptom onset, while relying on static educational and vocational attainments to retroactively estimate pre-morbid baseline. Implementing broadly accessible, self-administered longitudinal tracking of neurological domains, including assessment of higher cortical functions, could bridge this information gap, enabling clinical neurology to transition from a reactive framework to a proactive paradigm.

Digital biomarkers collected via ubiquitous devices such as smartphones or smartwatches offer a scalable approach to filling this monitoring void. Consumer hardware and software ecosystems already deploy applications that track cardiovascular health, physical activity, and athletic performance both passively and actively. Although passive monitoring offers high convenience, its unconstrained environment means that inferred anomalies (e.g., irregular heart rhythms or sudden falls), typically require active validation steps, such as a user-initiated electrocardiogram at rest or a confirmation prompt following a potential fall. While passive digital monitoring will undoubtedly continue to expand in breadth and fidelity, episodic, active, and highly structured testing will remain essential for the longitudinal assessment of targeted neurological domains.

We initially developed NeuFun-TS as an internal research tool to complement clinician-derived neurological examinations, increasing both the sensitivity and accuracy of identifying, quantifying, and longitudinally tracking neurological and cognitive deficits within our natural history protocol of neuroimmunological diseases and investigator-initiated clinical trials. A primary limitation of the bedside neurological examination that we sought to address with NeuFun-TS digital biomarkers is the comprehensive assessment of cognitive function. Here, we present findings for the fourth higher cognitive function test implemented in NeuFun-TS, building upon our previous validation of digitized SDMT (Pham et al., 2021) and verbal and spatial memory test (Kosa et al., 2026).

We prioritized implementing a digital motor sequencing paradigm for three reasons. First, motor praxis relies on complex, multisynaptic, bilateral frontoparietal and cortico-subcortical networks, heightening sensitivity to subclinical functional decline across disparate brain regions that often escape standard bedside neurological examinations. Second, intact motor sequencing is essential for real-world functioning, directly underlying everyday activities of daily living and vocational performance. Third, the task is intuitive across the lifespan and well-suited for automated administration and scoring on a smartphone.

In this study, we validated successful digital adaptation of the Luria FEP paradigm within NeuFun-TS across a prospective cohort of 296 participants including healthy donors and pwMS spanning the full clinical spectrum. The task yielded seven distinct digital biomarkers covering four dissociable motor subprocesses. Notably, six of these biomarkers significantly differentiated disease sub-cohorts along a consistent ordinal severity gradient (HD < RR-MS < P-MS). Furthermore, a parsimonious two-biomarker composite (MSC-2) demonstrated strong discrimination, achieving a concordance index of 0.87 for ordinal severity classification. These motor sequencing digital biomarkers exhibited robust convergent validity, correlating significantly with structural MRI metrics of CNS tissue damage, targeted subpanels of the bedside neurological examination, and independently derived cognitive measures.

The requirement to maintain a three-element motor program in working memory and execute it in precise temporal sequence under time pressure distinguishes our task from existing digital motor assessments. The Draw a Shape Test (Graves, Ganzetti, et al., 2023), Pinching Test (Graves, Elantkowski, et al., 2023), passive keystroke dynamics (Lam et al., 2021), Multiple Sclerosis Performance Test (Rao et al., 2020), as well as our own NeuFun-TS Tapping, Balloon-popping and Level tests (Boukhvalova et al., 2019; Boukhvalova et al., 2018) primarily evaluate downstream motor execution, including dexterity, speed, and movement accuracy. However, they do not demand the sequential ordering of distinct motor acts that characterizes the cognitive control of motor programming and dynamic praxis. Our paradigm addresses this critical gap by explicitly requiring the temporal serialization of motor programs, thereby allowing a granular decomposition into planning versus execution subprocesses that existing digital tools cannot capture.

This temporal decomposition revealed a fundamental dissociation: reaction time was independent of shape complexity, whereas drawing duration varied seven-fold across shape types. This confirms that inter-shape pauses reflect cognitive-motor planning, whereas active drawing reflects motor execution. The clinical utility of this distinction was demonstrated by speed-accuracy tradeoff analyses: patients with SP-MS predominantly exhibited coupled slow-and-inaccurate performance (84% in this quadrant), whereas those with PP-MS demonstrated a higher frequency of slow-but-accurate compensatory behavior (19% vs. 11% in SP-MS). This divergence aligns with the relative classic presentation of preferential motoric and infratentorial damage in PP-MS, which preserves cognitive motor programming, versus the diffuse cerebral pathology characteristic of SP-MS that impairs both planning and execution domains. Crucially, this pathophysiological distinction remains invisible to composite or single-metric digital biomarkers, emerging only through granular subprocess decomposition.

Finally, the convergent validity with NeuFun-TS SDMT and spatial memory biomarkers (rho = 0.49-0.61 for composites) demonstrates within-platform cross-task consistency while confirming that motor sequencing captures additional variance beyond what SDMT and memory tasks provide.

Several limitations warrant consideration. First, while test-retest reliability for MET and MSD was good (ICC = 0.74 and 0.68, respectively), accuracy measures exhibited lower reliability due to floor effects, limiting their utility for individual-level longitudinal tracking. Second, our HD cohort was younger than the MS groups; although biomarker differences persisted after adjusting for age in ordinal regression models, age-matched prospective validation would strengthen inference. Third, the current implementation was deployed on dedicated Google Pixel devices rather than participants’ personal smartphones, constraining immediate scalability. Conversely, expanding testing to heterogeneous consumer devices risks introducing algorithmic scoring bias driven by hardware-specific variability, such as screen aspect ratios or touch-sampling rates. Thus, when deployed as a software-as-a-medical-device (SaMD), each hardware-software pair may require distinct technical calibration. Fourth, longitudinal sensitivity for detecting true disease progression or therapeutic response remains to be established in prospective studies. To address this, we have modified the motor sequencing paradigm to yield a randomized, balanced stimulus composition (containing exactly one dot, line, and circle [DLC], with randomized temporal ordering and stroke directionality). Our sensitivity analysis comparing DLC-only trials to the full task battery (Supplementary Table S6) confirms that simplifying to balanced compositions eliminates the need for difficulty normalization while fully preserving diagnostic discrimination.

In conclusion, bridging the unmonitored gap in adult neurology requires scalable tools capable of capturing granular cognitive-motor dynamics before irreversible disability occurs. By digitizing the Luria FEP paradigm within NeuFun-TS, we demonstrate that unsupervised, smartphone-based motor sequencing provides robust digital biomarkers of CNS structural damage and disease severity in pwMS. Crucially, decomposing performance into planning versus execution subprocesses uncovers distinct speed-accuracy compensatory phenotypes in SP-MS versus PP-MS that are masked by conventional composite metrics. That a parsimonious composite of just two digital biomarkers achieves near-optimal clinical classification underscores the feasibility of incorporating targeted motor sequencing into routine practice, offering a high-yield, low-burden framework for proactive, precision neurological monitoring across the lifespan.

## Materials and Methods

### Study design and subjects

We conducted a prospective, observational study under two NIH protocols: NCT00794352 (“Comprehensive Multimodal Analysis of Neuroimmunological Diseases of the Central Nervous System [CNS]”) and NCT03109288 (“TRAP-MS: Targeting Residual Activity by Precision, Biomarker-Guided Combination Therapies of MS”) to evaluate a novel smartphone-based motor sequencing task. The Institutional Review Board of the NIH approved both protocols, and all participants provided written or electronic informed consent before any study procedures began. Data collection spanned from April 1, 2021 (the date the motor sequencing module launched) through June 11, 2026; every subject with at least one valid motor sequencing assessment in that interval was included.

Participants fell into three cohorts based on how and where they completed the tests. Cohort 1 consisted of HD who consented to the “smartphone only cohort” and performed all tasks unsupervised on investigator-provided smartphones. Cohort 2 included subjects who completed the motor sequencing test in person at the NIH Clinical Center, typically within one day of a full neurological evaluation and brain MRI. Cohort 3 comprised a subset of Cohort 2 participants who opted to continue home testing at their convenience. The results from all 3 cohorts were streamed to secured database under alphanumeric code. Data download, pre-processing and quality assessment was performed in blinded fashion.

Following unblinding of diagnostic categories, our analytic cohort comprised 296 participants: 34 HD, 101 RR-MS, 55 SP-MS, 52 PP-MS, 25 NIND, 21 OIND, and 8 CIS/RIS. A total of 3,278 trials were collected across all sessions. The primary analyses focused on the MS and HD groups (241 participants, 479 first-trial patient-hand observations); the NIND, OIND, and CIS/RIS groups were reserved for discriminant validity analyses reported alongside the primary group separation results. Participant demographics are detailed in Table 1 and Supplementary Figure S1.

### Clinical outcomes

Neurological exams were recorded in real time via the NeurEx App (Kosa et al., 2018) on iPads or desktop computers. The NeurEx App consists of 17 pages, each corresponding to a domain of the neurological examination where a clinician documents identified deficits using touch and swipe gestures on homunculus-like representations. The App algorithmically translates the documented neurological examination into traditional disability scales including the EDSS (ordinal scale from 0-10) (Kurtzke, 1983), the SNRS (Sipe et al., 1984), and associated Kurtzke Functional System Scores. Additionally, the NeurEx App scores all inputted data as a continuous disability scale from 0 (no disability) to a theoretical maximum of 1349. NeurEx subscores can be generated for any disability domain. CombiWISE, a composite disability scale optimized through machine learning, was derived from EDSS, SNRS, Timed 25-Foot Walk (T25FW) and non-dominant hand Nine-Hole Peg Test (9HPT) (Kosa et al., 2016). All prospectively-acquired clinical data are quality controlled weekly, after which the database inputs are locked to prevent future modifications.

For this study, we extracted NeurEx panel scores representing cognition, vision, upper extremities strength, pyramidal and cerebellar dysfuntion. We matched clinical assessments to NeuFun-TS trials within a 7-day window.

We collected SDMT scores through the validated smartphone module (Pham et al., 2021) and matched spatial memory composite scores from the validated spatial memory test (Kosa et al., 2026) on the same testing day.

### Imaging outcomes

Clinic-associated brain MRIs extending to the upper cervical spinal cord C5 level were scored using the COMRIS framework, a semi-quantitative grading system as previously described (Kosa et al., 2015). We used seven COMRIS-derived outcomes organized into three tiers: global CNS Tissue Destruction (COMRIS-CTD), supratentorial measures (supratentorial brain atrophy score, supratentorial T2 lesion load score), and infratentorial atrophy measures (infratentorial brain atrophy composite, brainstem atrophy, cerebellum atrophy, and medulla atrophy). The infratentorial brain atrophy composite was computed as the sum of brainstem, cerebellum, and medulla atrophy scores.

All scores are ordinal, derived from clinician-graded semi-quantitative ratings that occurs prospectively, within 5 days of MRI collection, following published scoring rules (Kosa et al., 2015). After quality control, the data are locked in the research database to prevent further modification.

### Software architecture and task protocol

The motor sequencing task was developed as a component of the Neurological Functional Test Suite (NeuFun-TS, Figure 1a), a growing collection of self-administered smartphone tests designed to assess distinct neurological domains within a single platform (Boukhvalova et al., 2019; Boukhvalova et al., 2018; Calcagni et al., 2025; Jin et al., 2024; Kosa et al., 2026; Messan et al., 2021; Pham et al., 2021).

The motor sequencing module was developed in Kotlin and Java using Android Studio and optimized for Google Pixel XL and Pixel 2 XL devices. The interface utilizes a standardized color palette designed to accommodate individuals with color vision deficiencies or those experiencing red color desaturation. The trial protocol consists of an initial training phase followed by an active test. Participants first practice drawing seven distinct symbol variations (two circles, two horizontal lines, two vertical lines, and one dot) with prescribed directionalities (Figure 1b-c). After achieving proficiency, they complete three guided practice runs of a target three-symbol sequence. The active test consists of a 30-second timed trial during which participants must reproduce the target sequence from memory as rapidly and accurately as possible. Real-time visual feedback confirms correct entries or prompts corrections (Figure 1d). This procedure is performed with the left hand first, then repeated with the right hand using a novel sequence.

### Data acquisition and logging

For each discrete drawing event, the application logged the expected shape type, the received shape type, the automated recognition status, and the corresponding raw touch coordinates (pointsDrawn). To capture fine-grained kinetic profiles, these raw (x, y) touch coordinates were continuously sampled at the device’s native display refresh rate of approximately 60 Hz (yielding one data sample every ∼16.7 ms), with highly precise relative timestamps appended to each recorded coordinate point.

All raw NeuFun-TS outputs streamed in real time to a private database hosted on Google Firebase. We deployed custom extraction scripts to pull JSON logs for each trial. All raw JSON logs are linked to anonymized subject IDs within Firebase and transferred securely to National Institute of Allergy and Infectious Diseases (NIAID) encrypted servers for scoring, quality control, and integration with clinical and imaging data. After integrity checks, we computed trial scores according to algorithms detailed in the Results.

### Digital biomarkers

We extracted seven digital biomarkers from each valid trial, spanning four primary domains of motor sequencing performance: 1. sequencing throughput, 2. speed, 3. motor execution, and 4. accuracy (Figure 1e). We present the comprehensive details of biomarker derivation, including the validation profiles for our timing decomposition and difficulty normalization, in the Results section.

We quantified the *Motor Sequencing Deficit (MSD)* as the deviation from a theoretical maximum of difficulty-normalized sequence throughput using the following model:

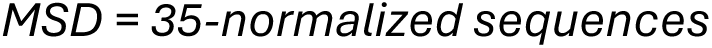

where higher values indicate greater impairment. We derived the theoretical maximum of 35 sequences from the fastest physiologically plausible component times within our dataset (1st-percentile reaction time = 122 ms, dot drawing = 37 ms, line drawing = 116 ms, and circle drawing = 318 ms), which collectively yielded an optimal baseline of 837 ms per sequence composed of dot, line, and circle, within the 30,000 ms trial window.

We decomposed the speed domain into three distinct kinematic components:

1. The *Individualized Sensory-Motor Processing Threshold (iSMPT),* which we defined as the minimum valid reaction time recorded within a trial.
2. The *Median Reaction Time (Median RT)*, which reflects combined cognitive and motor initiation latency.
3. The planning latency above threshold, termed the *adjusted Reaction Time Latency (RTLa)*, which we calculated as:

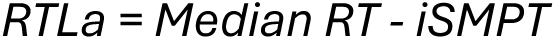

We quantified the *Motor Execution Time (MET)* as the median drawing duration following our shape-complexity adjustment. Finally, we partitioned accuracy into sequencing errors captured by the *Motor Sequencing Index (MSI*; log_10_(incorrect shapes / recognized shapes)), and shape recognition errors, captured by the *Shape Drawing Index (SDI*; log_10_ (unrecognized shapes / total shapes)).

We computed MET from all recognized drawing events with valid slope-adjusted durations, requiring at least two distinct shape categories per trial for a valid slope estimate. This approach maximized the number of contributing events per trial while the per-trial OLS slope adjustment ensured that shape-complexity bias was removed regardless of the specific shape composition drawn.

### Data processing and event classification

We processed the data through a multi-step analytical pipeline. First, we parsed event-level JSON logs for each trial. We classified each discrete drawing event into one of three categories based on sequence progression patterns: *correct*, *incorrect* (where the algorithm recognized the shape, but it was not intended shape in the sequence), or *unrecognized* (where the participant attempted the shape, but it failed to meet the detection threshold of the automated shape classification algorithm; Supplementary Figure S2). To derive the two log-transformed accuracy biomarkers (MSI and SDI), we adjusted trials yielding zero errors with a fixed floor value of 0.001 prior to log_10_ transformation to prevent undefined mathematical values.

### Kinematic decomposition and complexity adjustment

We decomposed the inter-shape interval using raw touchscreen coordinate trajectories (*pointsDrawn*) into two primary kinematic phases: *reaction time (RT*; the finger-up-to-finger-down interval reflecting motor planning and initiation) and drawing duration (the finger-down-to-finger-up interval reflecting motor execution). To isolate execution mechanics from geometric variations, we adjusted drawing durations for shape complexity using a per-trial *OLS* regression slope. We assigned non-equidistant complexity weights empirically derived from healthy control baseline data (dot = 0, line = 0.38, circle = 1.00).

The complexity adjustment isolated the baseline execution speed of the trial according to the following model:

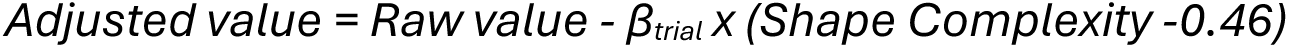

where 0.46 represents the mean complexity across all three shape categories, and *β_trial_* is the calculated trial-specific OLS slope. The adjustment removed the shape complexity effect while preserving the trial’s overall speed level. We applied stringent kinematic data filtering rules to eliminate artifactual data: we discarded RTs below 50 ms (physiologically implausible for voluntary motor responses) or above 5000 ms (indicative of task disengagement) and excluded drawing durations exceeding 3000 ms (indicative of pauses or hesitations unrelated to continuous motor execution).

### Sequence identification and normalization

We extracted the target three-shape sequence for each trial from the Firebase metadata (*sumData.sequenceExpected*). While the design concept intended to generate pseudo-random sequences that contain 3 different shapes (i.e., dot, line and circle), randomizing their sequence and directionality (i.e., left-to-right and right-to-left for horizonal line, top-to-button and bottom to top for vertical line, and clockwise- and counter-clockwise for circle) the horizontal and vertical lines were interpreted as separate shapes. This led to three structural composition categories that differed in the degree of drawing length: *Dot+Line+Line (DLL) < Dot+Line+Circle (DLC)< Circle+Line+Line (CLL)*. Within the DLL and CLL categories, the line elements had always different orientations (one horizontal and one vertical), ensuring that every sequence position demanded a distinct motor program.

A non-greedy string-matching algorithm detected complete sequences within each trial. We defined a complete sequence as three consecutive correct shapes that fully represented all three distinct base shape types. Once the algorithm identified a valid triplet, it advanced past the sequence boundaries to prevent overlapping counts (Supplementary Figure S2). Finally, we computed trial-specific difficulty factors based on the mathematically expected completion time for each unique sequence composition, enabling the normalization of raw sequence counts across the three composition categories.

### Quality control and exclusion criteria

We applied trial-level *quality control (QC)* using a three-tier classification system (Supplementary Figure S3a).

Because Cohort 1 consisted of self-declared HD who consented to the “smartphone only cohort” via the NeuFun-TS App and performed all tasks unsupervised, we labeled trials with floor performance (zero complete sequences) or fewer than five valid reaction times (RTs) in Cohort 1 as “*fail*” tier. In contrast, all first (and remaining clinic-associated) NeuFun-TS trials of pwMS were collected in clinic under supervision of NIH investigators (Cohort 2), who eliminating technical outliers immediately and asked the patient to repeat failed run. Therefore, statistical outliers from Cohort 2 represent genuine phenotypic expressions of disease biology.

The “*suspect*” tier flagged any trial that met at least one of five distinct statistical criteria: (1) an extreme RT exceeding three times the group *median absolute deviation (MAD)*; (2) a within-trial *coefficient of variation (CV)* exceeding 1.5; (3) a concurrent MAD-based outlier designation on two or more biomarkers; (4) a within-subject outlier exceeding two *standard deviations (SD)* from the participant’s own mean across a minimum of three trials; or (5) a learning-curve violation within the HD cohort, where the first trial yielded less than half the performance of subsequent attempts. When MS participants met these floor or insufficiency criteria, we assigned their trials to the “*suspect*” tier rather than the “*fail*” tier. We classified all remaining trials as “*pass*”.

To further safeguard data integrity, we applied a secondary, subject-level exclusion screen. We classified HD participants who recorded zero “*pass*” trials across all attempts as systematically disengaged and consequently promoted all of their trials to “*fail*” status. Ultimately, we excluded all “fail” trials from our final analytical datasets. Conversely, we retained “*suspect*” trials in all primary analyses; a subsequent sensitivity analysis confirmed that excluding these suspect trials did not alter our group separation metrics (see Results).

### Motor sequencing composite

To develop a composite biomarker of motor sequencing impairment, termed the Motor Sequencing Composite (MSC), we applied ordinal logistic regression (OLR) with clinical severity ordered as HD<RR-MS<P-MS, where P-MS combined SP-MS and PP-MS phenotypes. This approach follows the same structural framework we validated in our prior spatial memory study, demonstrating that aggregating multiple digital biomarkers into a single composite metric amplifies the clinical signal relative to any single constituent biomarker.

We randomly partitioned the participant pool into a training set (358 observations from 180 patients) and a validation set (185 observations from 94 patients), stratifying the split by clinical group, age, sex, and total NeurEx score. To prevent data leakage, we assigned both hands from an individual patient to the same analytical set. Because of their limited sample size, we replicated HD observations across both the training and validation sets. All seven biomarkers entered the full model; however, RTLa was automatically excluded during fitting due to perfect collinearity with Median RT and iSMPT (RTLa = Median RT − iSMPT), yielding six effective predictors in the full model (MSC-6: MSD, Median RT, iSMPT, MET, MSI, and SDI). We then applied bidirectional stepwise Akaike Information Criterion (AIC) selection to isolate the most parsimonious model, which ultimately retained two final predictors (MSC-2: MSD and Median RT). We fitted both models using the proportional-odds function and evaluated their performance within the independent validation cohort. Specifically, we assessed model performance using the concordance index, three-class exact accuracy, adjacent accuracy (which permitted misclassification by a single ordinal level), and confusion matrices.

### Trial selection framework

To ensure statistical independence and cross-analysis comparability, we tailored our trial selection criteria to each specific analytical question. For cross-sectional group separation, we selected the first trial per patient-hand to provide independent observations unconfounded by practice effects or temporal changes. For clinical validation (correlations with NeurEx, MRI, and functional measures), we isolated the first trial completed within 7 days of a clinic visit, establishing strict temporal proximity between the smartphone assessment and the clinical examination. For the MSC, we utilized the first trial per patient-hand and applied a 70/30 stratified train/validate split to prevent repeated-measures violations.

To maintain comparable measurement conditions across cohorts during reliability and learning analyses, we instituted strict temporal constraints. HD (Cohort 1) completed testing sessions approximately every day (median inter-trial interval of approximately 1 day), whereas pwMS tested primarily during periodic clinic visits (Cohort 2; median inter-trial interval spanning several months). Without these temporal constraints, analyzing the “first 10 trials” would capture a 10-day span for HD participants but potentially span several years for MS patients, heavily confounding short-term practice effects with long-term disease progression. We therefore evaluated test-retest reliability using the first three trials per patient-hand completed within a strict 90-day window, capturing the stability of the digital measurement itself rather than biological trait changes. We selected this three-trial, 90-day threshold because MS patients can realistically achieve it through combined clinic-based and home testing, while the brief interval ensures that disease progression remains negligible.

### Mathematical modeling and statistical methods

We applied a log_10_ transformation to any dataset displaying a non-normal distribution. To evaluate associations between continuous variables, we calculated Spearman’s rank-order correlation coefficient (rho). For linear regression models, we report the coefficient of determination (R^2^), the corresponding p-value, and the total sample size (N). Where appropriate, we computed Lin’s concordance correlation coefficient (CCC) to assess agreement between measured and predicted outcomes directly. We evaluated intergroup differences using either the Wilcoxon rank-sum test for two-group comparisons or the Kruskal-Wallis test followed by post-hoc pairwise Wilcoxon rank-sum comparisons for multi-group analyses. We present raw p-values throughout unless we explicitly note an adjustment for multiple comparisons.

We evaluated the test-retest reliability of our longitudinal measurements by calculating the intraclass correlation coefficient (ICC form 3,1: two-way consistency, single measures) across the first three trials per patient-hand completed within a 90-day window. To characterize performance trends and learning effects over time, we constructed linear mixed-effects models specifying a trial number-by-group interaction term alongside random intercepts for each unique patient-hand. Where applicable, we computed 95% confidence intervals via bootstrapping with 2,000 iterations, resampling data at the individual participant level. We performed all statistical analyses and data modeling within R version 4.5.1(RCoreTeam, 2025).

### Luria-derived analyses

Motivated by the Luria FEP paradigm (Dubois et al., 2000; Luria, 1966), we performed six event-level analyses to characterize the *qualitative* nature of motor sequencing deficits. We computed per-patient descriptive measures (including error classifications and transition matrices) from the first 10 trials per patient-hand to establish a sufficiently robust event pool. For all intergroup comparisons, we utilized single first-trial data points (one observation per patient-hand) and evaluated them using Kruskal-Wallis tests. These six core analyses evaluated:

1. Error taxonomy and the perseveration index. We classified each sequencing error (WRONG_SHAPE) based on its relationship to the expected sequence: *perseverative* (the drawn shape matched the previously expected target, indicating repetition of a prior motor program), *anticipatory* (the drawn shape matched the next expected target, indicating premature sequence advancement), or *other*. We then computed a trial-level perseveration index:

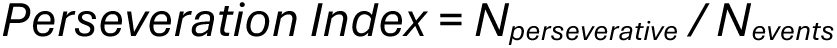

where N_perseverative_ is the number of perseverative errors and N_events_ is the total number of drawing events in the trial.

2. Switch cost. For each drawing event with a valid RT, we classified the shape transition as *Repeat* (same shape category as previous event) or *Switch* (different shape category). We computed the trial-level switch cost as:

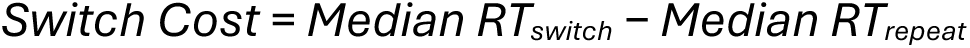

where positive values indicate additional processing time for transitions between distinct motor programs.

3. Within-trial deceleration. We divided each trial’s valid drawing events into positional tertiles (Early, Middle, Late) and computed:

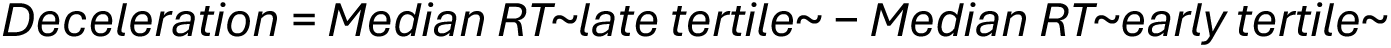

where positive values indicate slowing across the 30-second trial (micro-fatigue) and negative values indicate warm-up acceleration.

4. Error clustering. We identified consecutive runs of sequencing errors within each trial and computed the mean error run length. A mean run length approaching 1.0 indicates isolated errors; values substantially above 1.0 indicate cascading failures where errors compound.
5. Speed-accuracy quadrant distributions. We plotted each patient-hand in a two-dimensional space (Median RT × MSI), defined quadrants relative to HD medians, and evaluated group differences in quadrant occupancy via chi-squared tests.
6. Complexity-by-group interactions on drawing duration. We used the per-trial OLS complexity slope (β_trial_, defined above) as the outcome measure, assessing whether progressive MS patients pay a disproportionate motor execution cost for complex shapes.

For the perseveration analysis, we restricted the sample to patient-hands that committed at least one sequencing error (N = 339). We applied this constraint because a perseveration index of zero derived from a zero-error trial reflects an absence of data rather than low perseveration, whereas a zero index from a trial containing errors provides informative phenotypic variation. For the speed-accuracy dissociation analysis, we excluded patient-hands with a MSI at the floor value of -3 (signifying zero sequencing errors; N = 339). We omitted these individuals because they did not participate in the standard speed-accuracy tradeoff; their accuracy remained perfect by definition, independent of execution speed.

### Sensitivity analyses

We conducted four sensitivity analyses to evaluate the robustness of our methodological choices.

First, to determine if a more stringent QC filter improves group separation, we compared our primary analysis (which retained all suspect trials) against a targeted filter designed to exclude only trials with unambiguous measurement artifacts. This targeted filter isolated trials meeting any of three criteria: within-subject outliers (where a patient’s longitudinal history indicated highly atypical performance), a high coefficient of variation (where erratic within-trial timing suggested participant disengagement or technical issues), or convergent evidence from three or more simultaneous flags. We designed this targeted filter to remove technical noise while preserving genuine disease biology, as single-criterion flags, such as a MAD outlier or extreme RT disproportionately capture true motor impairment in progressive MS rather than technical error.

Second, to assess whether our shape-complexity adjustment and difficulty normalization protocols provided incremental value beyond naturally balanced trials, we compared two distinct analytical strategies. We contrasted a “DLC-only” approach (i.e., restricted to trials containing exactly one dot, one line, and one circle, which required no mathematical adjustment) against our full analytical pipeline, which applied slope adjustment and difficulty normalization across all trial compositions. We utilized biomarkers requiring no adjustment (i.e., Median RT, iSMPT, RTLa, MSI, SDI) as internal controls. Equivalent performance across these control metrics confirmed a comparable patient composition between the datasets, allowing us to attribute any divergent performance in the adjusted biomarkers (i.e., MSD, MET) directly to the modeling methodology rather than underlying sample differences.

Third, to evaluate whether our biomarker sensitivity uniquely reflected MS-specific pathology or merely captured generic neurological illness, we extended the analysis to include cohorts with non-inflammatory neurological disease (NIND), other inflammatory neurological disease (OIND), and clinically or radiologically isolated syndromes (CIS/RIS).

## Data Availability

All data produced are available online at Github repository upon manuscript acceptance by a journal.

https://github.com/Bielekova-Lab/neufun-motor-sequencing

## Abbreviations

9HPT: Nine-Hole Peg Test
AIC: Akaike Information Criterion
AUROC: Area Under the Receiver Operating Curve
CCC: concordance correlation coefficient
CI: confidence interval
CIS/RIS: clinically isolated syndrome / radiologically isolated syndrome
CNS: central nervous system
CombiWISE: Combinatorial Weight-adjusted Disability Scale
COMRIS: Combinatorial MRI Scale
COMRIS-CTD: COMRIS-CNS Tissue Destruction
CV: coefficient of variation
DLC: Dot-Line-Circle (sequence composition)
DLL: Dot-Line-Line (sequence composition)
DynamicSRLa: Dynamic adjusted Spatial Response Latency (from NeuFun-TS spatial memory test)
CLL: Circle-Line-Line (sequence composition)
EDSS: Expanded Disability Status Scale
FEP: fist-edge-palm (Luria task)
fMRI: functional magnetic resonance imaging
HD: healthy donors
ICC: intraclass correlation coefficient
IQR: interquartile range
iSMPT: Individualized Sensory-Motor Processing Threshold
iSMPT-S: Individualized Sensory-Motor Processing Threshold - Spatial (from NeuFun-TS spatial memory test)
MAD: median absolute deviation
MET: Motor Execution Time
MRI: magnetic resonance imaging
MS: multiple sclerosis
MSC: Motor Sequencing Composite
MSC-2: Motor Sequencing Composite (2-predictor model: MSD, Median RT)
MSC-6: Motor Sequencing Composite (6-predictor model)
MSD: Motor Sequencing Deficit
MSI: Motor Sequencing Index
NeuFun-TS: Neurological Functional Test Suite
NIAID: National Institute of Allergy and Infectious Diseases
NIH: National Institutes of Health
NIND: non-inflammatory neurological disease
OIND: other inflammatory neurological disease
OLR: ordinal logistic regression
OLS: ordinary least squares
P-MS: progressive multiple sclerosis
PP-MS: primary-progressive multiple sclerosis
pwMS: people with multiple sclerosis
QC: quality control
RR-MS: relapsing-remitting multiple sclerosis
RT: reaction time
RTLa: adjusted Reaction Time Latency
SD: standard deviation
SDI: Shape Drawing Index
SDMT: Symbol Digit Modalities Test
SE: standard error
SMA: supplementary motor area
SNRS: Scripps Neurological Rating Scale
SP-MS: secondary-progressive multiple sclerosis
StaticSRLa: Static adjusted Spatial Response Latency (from NeuFun-TS spatial memory test)
T25FW: Timed 25-Foot Walk

## Author contribution

Conceptualization, B.B.; Methodology, P.K. and B.B.; Data curation, P.K.; Formal analysis, P.K. and B.B.; Investigation, A.M.A., M.K., Y.M., E.M., C.S., and B.B.; Visualization, P.K.; Writing – original draft, P.K. and B.B.; Writing – revision, All authors; Supervision, B.B.

## Data and code availability

All data and code used to generate the results in this manuscript will be publicly available in the project repository at https://github.com/Bielekova-Lab/neufun-motor-sequencing upon publication.

## Acknowledgments

We thank our patient care coordinator Michelle Woodland for her excellent care of our patients and our former postbaccalaureate fellows for help with data collection. We thank all research participants and patients’ caregivers for their time and effort.

