## Supplementary info for "Decomposing cognitive-motor planning from execution: smartphone motor sequencing provides scalable digital biomarkers of cognitive-motor function across central nervous system disorders"

### Supplementary Information

#### Supplementary Results

##### Difficulty normalization enables pooling of all trial compositions without sacrificing discrimination

The app sampled sequences from three composition categories (DLC, DLL, CLL), which varied in overall motor complexity. Because CLL sequences (two lines and a circle) inherently yielded fewer completions than DLL sequences (two lines and a dot) within the same 30-second window, raw sequence counts were not directly comparable across compositions. We developed a difficulty factor correction that eliminated this composition-dependent variance - after normalization, throughput was independent of sequence composition (Supplementary Figure S4a). However, the additional complexity of this normalization step motivated a design decision for future app versions: restricting sequences to balanced DLC compositions eliminates the need for difficulty normalization entirely. To validate this decision, we compared group separation (rank-biserial r, HD vs SP-MS) between DLC-only first trials (N = 219, requiring no difficulty normalization or complexity adjustment) and the full pipeline applying both corrections to all trial compositions (N = 479). We classified biomarkers into two categories: CONTROL biomarkers requiring no adjustment (Median RT, iSMPT, RTLa, MSI, SDI) served as internal controls for sample comparability, while ADJUSTED biomarkers (MSD, MET) tested whether the normalization steps preserved clinical signal. CONTROL biomarkers showed comparable effect sizes in both datasets (mean r: DLC = 0.59, Full = 0.61), confirming equivalent patient composition. For ADJUSTED biomarkers, DLC-only performed comparably: MSD was slightly lower (r = 0.63 vs 0.73), MET slightly higher (r = 0.53 vs 0.50; Supplementary Figure S4b). These results confirm that restricting sequences to balanced DLC compositions in future app versions will simplify the analytical pipeline without sacrificing discriminative validity, while eliminating the need for both difficulty normalization and shape-complexity slope adjustment of the MSD biomarker.

##### Seven digital biomarkers capture four dissociable motor subprocesses

The 7 x 7 Spearman correlation matrix (Supplementary Figure S5) revealed structured relationships among biomarkers. Speed measures (Median RT, iSMPT, RTLa) clustered together, as did accuracy measures (MSI, SDI). Motor Execution Time (MET) correlated moderately with speed but represented a partially independent motor execution dimension. Motor Sequencing Deficit (MSD) showed the broadest correlations, which is expected for an integrative throughput measure that depends on both speed and accuracy. This structure supports that the seven biomarkers, while derived from a single 30-second trial, tap into dissociable subprocesses of motor sequencing.

#### Targeted QC filter sensitivity analysis (Supplementary Figure S3)

To evaluate whether a more stringent QC approach could improve group separation, we compared the primary analysis against a targeted filter that excluded within-subject outliers, high CV trials, or trials meeting three or more other criteria simultaneously. The targeted filter removed 228 of 3,061 non-fail trials (7.4%), losing only 14 first-trial observations (N = 465, 240 patients vs primary N = 479, 241 patients). Mean absolute effect size (Wilcoxon r, HD vs SP-MS) was comparable between approaches (mean r = 0.413 vs 0.412 in the primary analysis), with four of seven biomarkers showing increased effect sizes, with the largest gain in iSMPT (+0.062). The targeted filter selectively removes measurement artifacts — erratic timing patterns and trials inconsistent with the patient's own longitudinal trajectory — while preserving the extreme but genuine motor impairment captured by single-flag criteria. These results validate the primary analysis decision to retain suspect trials, as the targeted filter produces negligible overall improvement in group discrimination despite additional sample loss.

#### Supplementary Tables

##### Supplementary Table S1. Group separation of seven digital biomarkers using first-trial data (N = 479 patient-hand observations).

**A: Biomarker distributions by group**

| Biomarker | HD | RR-MS | SP-MS | PP-MS | KW p | r (HD vs SP-MS) |
|---|---|---|---|---|---|---|
| MSD (score) | 23.9 [20.6, 27.0] | 27.1 [24.0, 30.0] | 30.9 [28.0, 33.0] | 30.0 [26.9, 33.0] | $1.1 \times 10^{-21}$ | 0.73 |
| Median RT (ms) | 351.0 [270.0, 537.0] | 513.0 [350.5, 673.0] | 854.5 [545.2, 1276.5] | 727.0 [455.0, 1182.0] | $8.0 \times 10^{-21}$ | 0.69 |
| iSMPT (ms) | 193.0 [152.0, 334.0] | 234.0 [177.0, 356.0] | 346.5 214.8, 555.8] | 309.0 [199.0, 556.0] | $2.0 \times 10^{-7}$ | 0.39 |
| RTLa (ms) | 152.0 [109.0, 193.0] | 222.0 [147.5, 312.0] | 380.0 [207.2, 764.1] | 304.5 [190.5, 479.5] | $3.3 \times 10^{-15}$ | 0.62 |
| MET (ms) | 276.5 [231.7, 375.6] | 352.9 [288.2, 448.9] | 431.1 [332.8, 572.8] | 424.7 [331.0, 576.4] | $4.9 \times 10^{-11}$ | 0.5 |
| MSI (log10) | -1.4 [-3.0, -1.1] | -1.4 [-3.0, -1.1] | -1.0 [-2.7, -0.6] | -1.2 [-3.0, -0.7] | $8.9 \times 10^{-5}$ | 0.36 |
| SDI (log10) | -3.0 [-3.0, -1.7] | -3.0 [-3.0, -1.5] | -3.0 [-3.0, -1.2] | -3.0 [-3.0, -1.3] | 0.14 | 0.14 |

**B: Pairwise comparisons (Holm-corrected Wilcoxon rank-sum tests)**

| Biomarker | HD vs RR-MS | HD vs SP-MS | HD vs PP-MS | RR-MS vs SP-MS | RR-MS vs PP-MS | SP-MS vs PP-MS |
|---|---|---|---|---|---|---|
| MSD | r = 0.29 * | r = 0.61 * | r = 0.54 * | r = 0.39 * | r = 0.29 * | NS |
| Median RT | r = 0.23 * | r = 0.57 * | r = 0.49 * | r = 0.42 * | r = 0.29 * | NS |
| iSMPT | NS | r = 0.33 * | r = 0.29 * | r = 0.25 * | r = 0.20 * | NS |
| RTLa | r = 0.25 * | r = 0.52 * | r = 0.46 * | r = 0.32 * | r = 0.23 * | NS |
| MET | r = 0.25 * | r = 0.42 * | r = 0.46 * | r = 0.20 * | r = 0.23 * | NS |
| MSI | NS | r = 0.30 * | NS | r = 0.22 * | NS | NS |
| SDI | NS | NS | NS | NS | NS | NS |

Values are median [IQR] in A. KW p = Kruskal-Wallis omnibus p-value. r = rank-biserial effect size. Panel B: * = $p < 0.05$ (Holm-corrected); NS = not significant. SP-MS vs PP-MS never reaches significance, consistent with overlapping disability profiles in progressive MS.

#### Supplementary Table S2. Test-retest reliability (ICC 3,1) of seven digital biomarkers.

**A: Primary analysis (first 3 trials within 90-day window)**

| Biomarker | ICC | 95% CI | N | p | Interpretation |
|---|---|---|---|---|---|
| MSD | 0.68 | [0.56, 0.78] | 65 | < 0.001 | Good |
| Median RT (ms) | 0.58 | [0.44, 0.71] | 65 | < 0.001 | Moderate |
| iSMPT (ms) | 0.60 | [0.46, 0.72] | 65 | < 0.001 | Good |
| RTLa (ms) | 0.31 | [0.15, 0.48] | 65 | < 0.001 | Fair |
| MET (ms) | 0.74 | [0.63, 0.82] | 65 | < 0.001 | Good |
| MSI | 0.16 | [0.01, 0.33] | 65 | 0.020 | Poor |
| SDI | 0.26 | [0.10, 0.43] | 65 | < 0.001 | Fair |

**B: Secondary analysis (up to 10 trials, unrestricted time span)**

| Biomarker | ICC | 95% CI | N | p |
|---|---|---|---|---|
| MSD | 0.75 | [0.66, 0.83] | 328 | < 0.001 |
| Median RT (ms) | 0.64 | [0.54, 0.75] | 328 | < 0.001 |
| iSMPT (ms) | 0.38 | [0.28, 0.52] | 328 | < 0.001 |
| RTLa (ms) | 0.46 | [0.35, 0.59] | 328 | < 0.001 |
| MET (ms) | 0.69 | [0.59, 0.79] | 328 | < 0.001 |
| MSI | 0.14 | [0.07, 0.25] | 328 | < 0.001 |
| SDI | 0.23 | [0.14, 0.35] | 328 | < 0.001 |

ICC(3,1): two-way consistency, single measures. A interpretation thresholds: Poor (< 0.40), Fair (0.40–0.59), Moderate (0.60–0.74), Good (≥ 0.75). The secondary analysis uses more trials over a longer span, increasing N but potentially confounding with disease progression and practice effects. MSD and MET remain the most reliable biomarkers across both analyses.

#### Supplementary Table S3. Sensitivity analysis: averaged trials versus single first trial.

| Biomarker | r (Average of 3 trials) | r (Single first trial) | Delta | Average better? |
|---|---|---|---|---|
| MSD | 0.73 | 0.73 | −0.002 | No |
| Median RT | 0.69 | 0.69 | +0.007 | Yes |
| iSMPT | 0.50 | 0.39 | +0.105 | Yes |
| RTLa | 0.55 | 0.62 | −0.071 | No |
| MET | 0.62 | 0.50 | +0.123 | Yes |
| MSI | 0.44 | 0.36 | +0.082 | Yes |
| SDI | 0.59 | 0.14 | +0.457 | Yes |

Effect sizes (rank-biserial r, HD vs SP-MS) comparing the mean of the first 3 trials within 90 days (N = 216 patient-hands) versus the single first trial (N = 479). Averaging improved discrimination in 5 of 7 biomarkers, with the largest gain for SDI (+0.46), confirming that repeat testing reduces measurement noise. RTLa and MSD showed minimal change, consistent with their already high single-trial reliability.

#### Supplementary Table S4. Ordinal logistic regression performance.

**A: Model coefficients (MSC-2: 2-predictor stepwise model)**

| Predictor | Coefficient |
|---|---|
| MSD | 0.156 |
| Median RT (ms) | $8.71 \times 10^{-4}$ |

**B: Validation performance (MSC-2)**

| Metric | Value |
|---|---|
| C-index (train) | 0.88 |
| C-index (test) | 0.868 |
| Exact accuracy (test) | 47.0% |
| Adjacent accuracy (test) | 93.0% |
| Spearman rho (test) | 0.381 |
| N (train) | 358 |
| N (test) | 185 |

**C: Univariate ordinal C-indices (individual biomarkers, validation set)**

| Biomarker | Coefficient | C-index (test) | AIC |
|---|---|---|---|
| MSD | 0.215 | 0.739 | 656 |
| RTLa (ms) | 0.00238 | 0.724 | 706 |
| Median RT (ms) | 0.00236 | 0.720 | 672 |
| MET (ms) | 0.00276 | 0.676 | 714 |
| iSMPT (ms) | 0.00265 | 0.626 | 711 |
| MSI | 0.217 | 0.610 | 747 |
| SDI | 0.299 | 0.524 | 744 |

Ordinal logistic regression with severity ordered as HD < RR-MS < P-MS. MSC-3 selected via bidirectional stepwise AIC from 6 candidate predictors. C-index generalizes AUROC to ordinal outcomes. Adjacent accuracy permits misclassification by one ordinal level. C demonstrates that the MSC-2 composite (C = 0.868) substantially outperforms any single biomarker (best individual: MSD, C = 0.739).

#### Supplementary Table S5. Summary of six Luria-derived analyses.

##### A: Statistical results

| Analysis | Test statistic | p | Result | Interpretation |
|---|---|---|---|---|
| Complexity × Group interaction | KW H = 227.2 | $5.6 \times 10^{-49}$ | Significant | MS groups show steeper drawing duration increase with shape complexity; motor execution capacity limitation |
| Error taxonomy (Perseveration Index) | KW H = 10.9 | 0.013 | Significant | SP-MS elevated perseverative errors (3x HD rate); validates digital Luria construct |
| Speed-accuracy dissociation | $\chi^2 = 77.8$ | $4.4 \times 10^{-13}$ | Significant | HD: speed/accuracy independent; SP-MS: coupled deficit (slow + inaccurate); PP-MS: compensates (slow, preserves accuracy) |
| Switch cost | KW H = 2.02 | 0.57 | Null | MS slowing is generalized across all transitions, not switching-specific |
| Within-trial deceleration | KW H = 0.97 | 0.81 | Null | All groups accelerate within trial; 30-s trial does not produce micro-fatigue |
| Error clustering | KW H = 1.46 | 0.69 | Null | Errors are isolated events (mean run ~1.2); no cascading failures |

##### B: Error taxonomy by group

| Group | Perseverative | Anticipatory | Other | Total errors |
|---|---|---|---|---|
| HD | 265 (36.4%) | 78 (10.7%) | 385 (52.9%) | 728 |
| RR-MS | 433 (35.0%) | 143 (11.6%) | 662 (53.5%) | 1238 |
| SP-MS | 327 (34.9%) | 132 (14.1%) | 478 (51.0%) | 937 |
| PP-MS | 263 (33.3%) | 80 (10.1%) | 447 (56.6%) | 790 |

All analyses use first-trial data (one observation per patient-hand). Kruskal-Wallis tests compare four groups (HD, RR-MS, SP-MS, PP-MS). Perseveration and speed-accuracy analyses restricted to patient-hands with at least one sequencing error (N = 339). B shows that the proportion of perseverative errors is stable across groups (33–36%), indicating MS increases error frequency without altering the qualitative error profile.

#### Supplementary Table S6. Sensitivity analysis: DLC-only versus full pipeline.

| Biomarker | Type | r (DLC-only) | r (Full pipeline) | DLC better? |
|---|---|---|---|---|
| MSD | ADJUSTED | 0.63 | 0.73 | No |
| MET | ADJUSTED | 0.53 | 0.50 | Yes |
| Median RT | CONTROL | 0.70 | 0.69 | Yes |
| iSMPT | CONTROL | 0.30 | 0.39 | No |
| RTLa | CONTROL | 0.68 | 0.62 | Yes |
| MSI | CONTROL | 0.58 | 0.61 | No |
| SDI | CONTROL | 0.67 | 0.73 | No |

Effect sizes (rank-biserial r, HD vs SP-MS) comparing DLC-only raw first trials versus the full pipeline with difficulty normalization and slope adjustment. N reflects the HD + SP-MS subset used for effect size computation (DLC: N = 69; Full: N = 175). ADJUSTED biomarkers require mathematical correction; CONTROL biomarkers need no adjustment and serve as internal controls for sample comparability. Comparable CONTROL effect sizes confirm equivalent patient composition between datasets.

#### Supplementary Figures

### Supplementary Figure S1

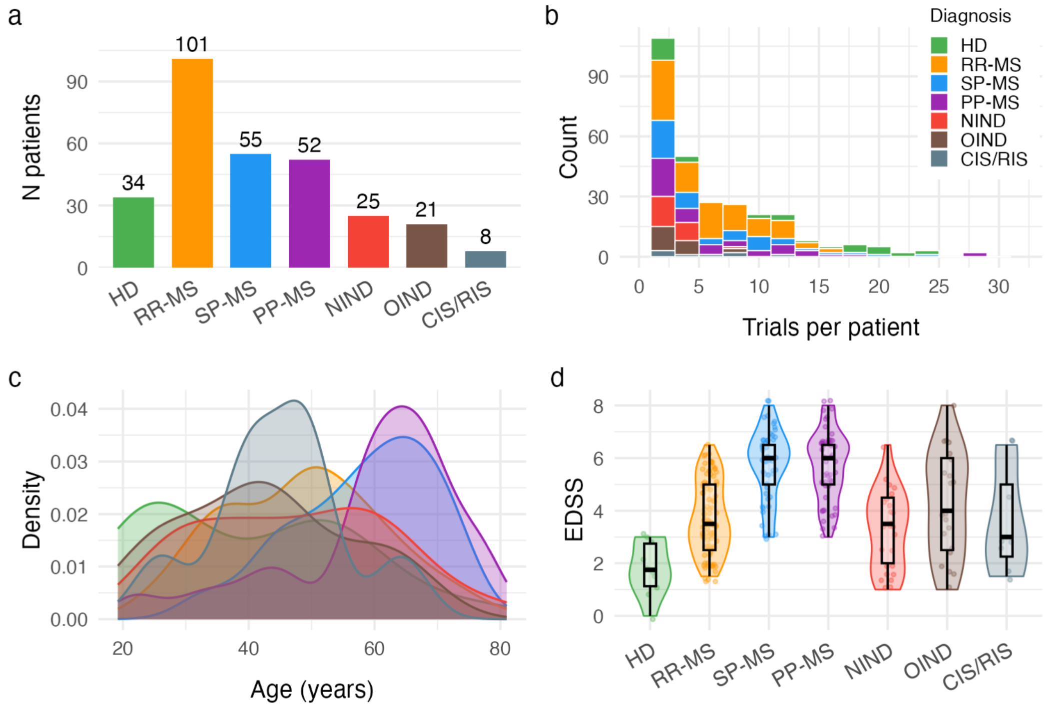


Supplementary Figure S1. Cohort overview.

(a) Bar chart of participant counts by diagnostic group. (b) Stacked histogram of trials per patient colored by diagnostic group. (c) Overlapping density plots of age distribution by group. (d) Violin plots showing EDSS distribution by group. HD = healthy donors; RR-MS = relapsing-remitting MS; SP-MS = secondary-progressive MS; PP-MS = primary-progressive MS; NIND = non-inflammatory neurological disease; OIND = other inflammatory neurological disease; CIS/RIS = clinically/radiologically isolated syndrome; EDSS = Expanded Disability Status Scale.

### Supplementary Figure S2

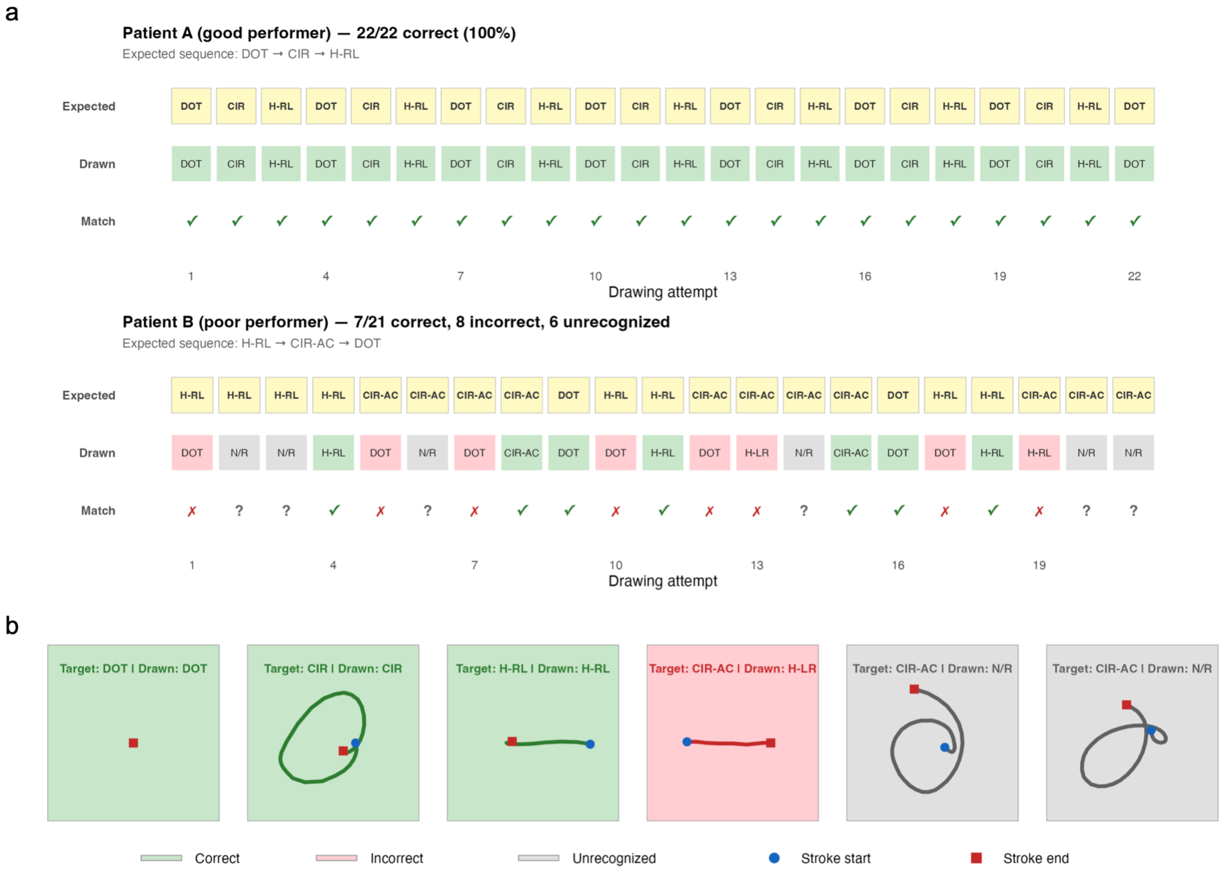


Supplementary Figure S2. Example patients: tape diagrams and finger trajectories.

(a) Tape diagrams showing the full trial sequence for two representative patients - a good performer (top) and a poor performer (bottom). For each drawing event, three rows display the Expected shape (yellow), the Drawn shape (green = correct, red = incorrect, gray = unrecognized), and the Match status. (b) Raw finger trajectory plots for six individual drawing events. Left panels (green background): correctly drawn shapes. Right panels: an incorrect shape (red background) and two unrecognized attempts (gray background). Blue circles mark stroke start; red squares mark stroke end.

### Supplementary Figure S3

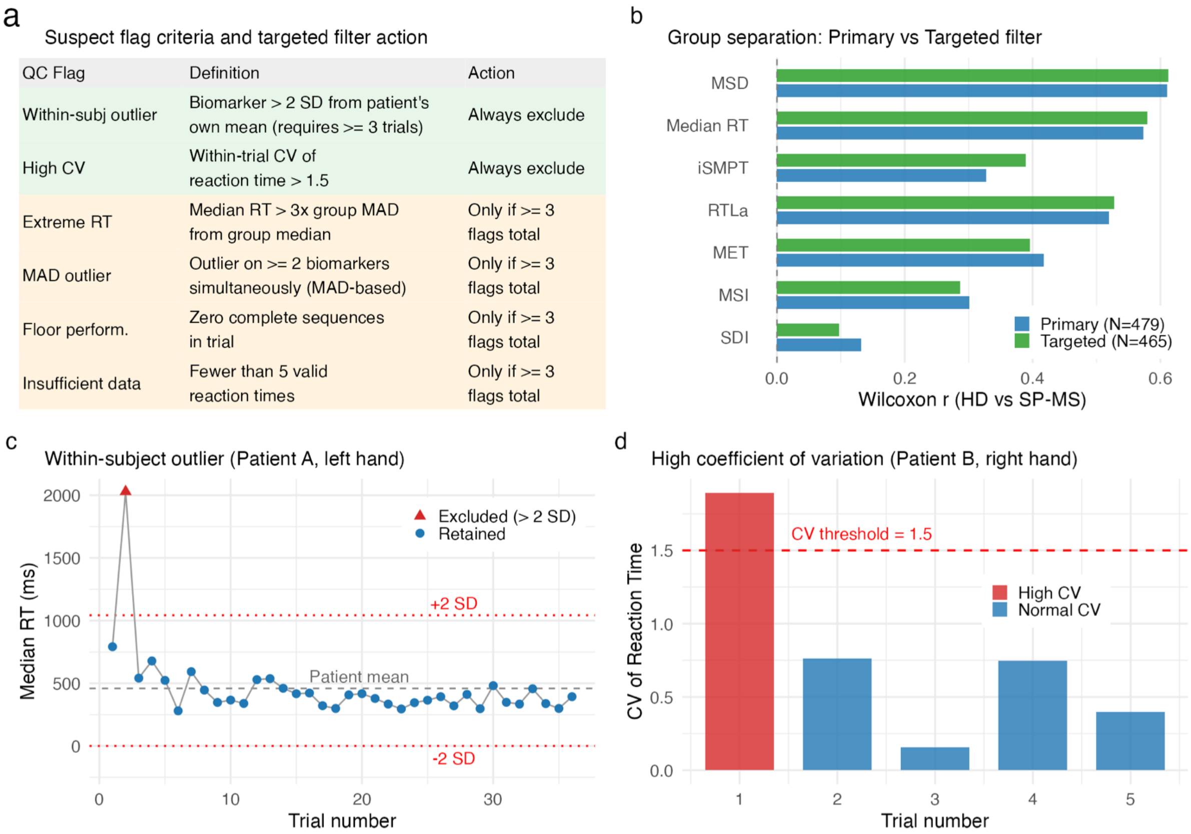


| QC Flag | Definition | Action |
|---|---|---|
| Within-subj outlier | Biomarker > 2 SD from patient's own mean (requires >= 3 trials) | Always exclude |
| High CV | Within-trial CV of reaction time > 1.5 | Always exclude |
| Extreme RT | Median RT > 3x group MAD from group median | Only if >= 3 flags total |
| MAD outlier | Outlier on >= 2 biomarkers simultaneously (MAD-based) | Only if >= 3 flags total |
| Floor perform. | Zero complete sequences in trial | Only if >= 3 flags total |
| Insufficient data | Fewer than 5 valid reaction times | Only if >= 3 flags total |

Supplementary Figure S3. Targeted QC filter validation.

(a) Table of suspect flag criteria and targeted filter actions. Two flags trigger automatic exclusion; four additional flags trigger exclusion only when 3 or more co-occur. (b) Horizontal bar chart comparing effect sizes (rank-biserial r, HD vs SP-MS) between the Primary filter (blue) and the Targeted filter (green). (c) Example of within-subject outlier detection: Median RT across trials for one patient, with mean and ± 2 SD thresholds shown. (d) Example of high coefficient of variation: within-trial CV of RT across trials, with CV = 1.5 threshold indicated. QC = quality control; SD = standard deviation; CV = coefficient of variation; RT = reaction time; MAD = median absolute deviation; HD = healthy donors; SP-MS = secondary-progressive MS.

### Supplementary Figure S4

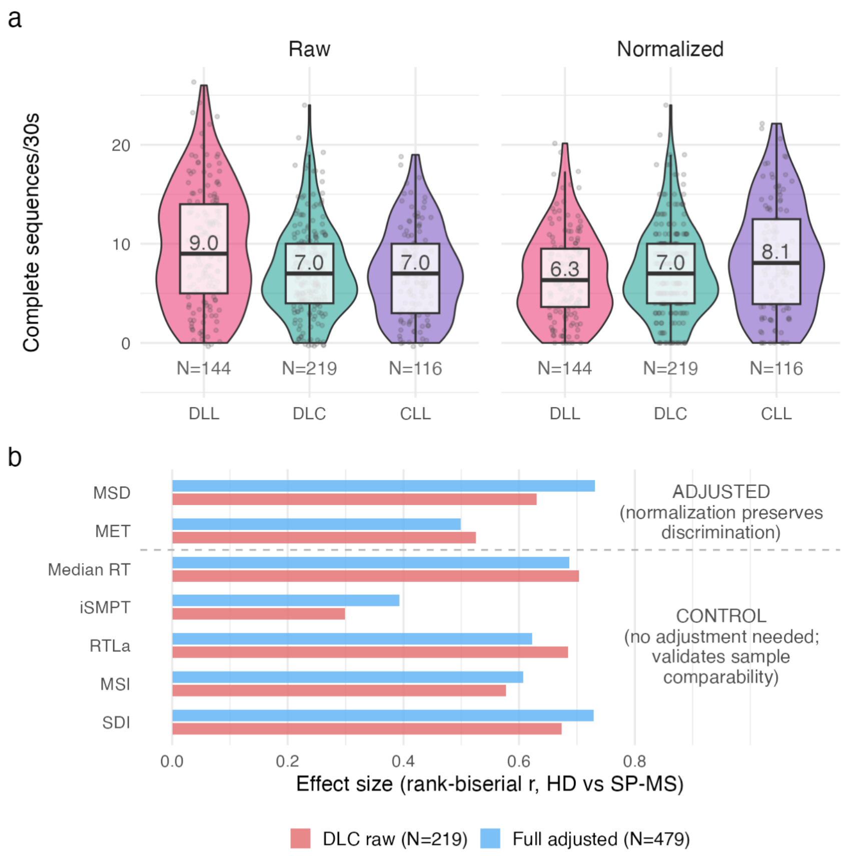


Supplementary Figure S4. Difficulty normalization enables pooling of all trial compositions.

(a) Violin plots comparing raw (left) and difficulty-normalized (right) complete sequence counts across three sequence compositions (DLL, DLC, CLL). Raw counts show a composition-dependent gradient that is equalized after normalization. (b) Horizontal bar chart comparing effect sizes (rank-biserial r, HD vs SP-MS) between DLC-only raw trials and the full adjusted pipeline for all seven biomarkers, divided into ADJUSTED (MSD, MET) and CONTROL (Median RT, iSMPT, RTLa, MSI, SDI) categories. DLL = Dot-Line-Line; DLC = Dot-Line-Circle; CLL = Circle-Line-Line; MSD = Motor Sequencing Deficit; MET = Motor Execution Time; Median RT = Median Reaction Time; iSMPT = individualized Sensory-Motor Processing Threshold; RTLa = Reaction Time Latency adjusted; MSI = Motor Sequencing Index; SDI = Shape Drawing Index; HD = healthy donors; SP-MS = secondary-progressive MS.

### Supplementary Figure S5

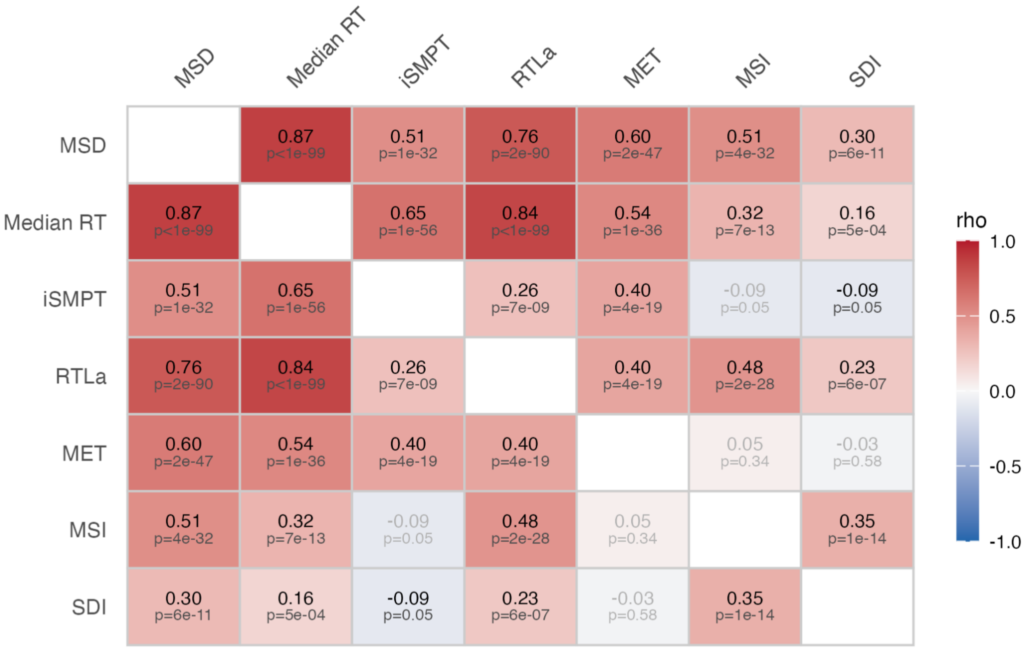


Supplementary Figure S5. Inter-biomarker correlation matrix.

Spearman rank correlation matrix (7 x 7) among all biomarkers computed from first-trial data. Each cell displays the correlation coefficient and FDR-corrected p-value. Color scale ranges from blue (negative) through white (zero) to red (positive). Speed measures cluster together, as do accuracy measures, while MET represents a partially independent motor execution dimension and MSD shows the broadest correlations consistent with an integrative throughput measure. MSD = Motor Sequencing Deficit; Median RT = Median Reaction Time; iSMPT = individualized Sensory-Motor Processing Threshold; RTLa = Reaction Time Latency adjusted; MET = Motor Execution Time; MSI = Motor Sequencing Index; SDI = Shape Drawing Index; FDR = false discovery rate.

### Supplementary Figure S6

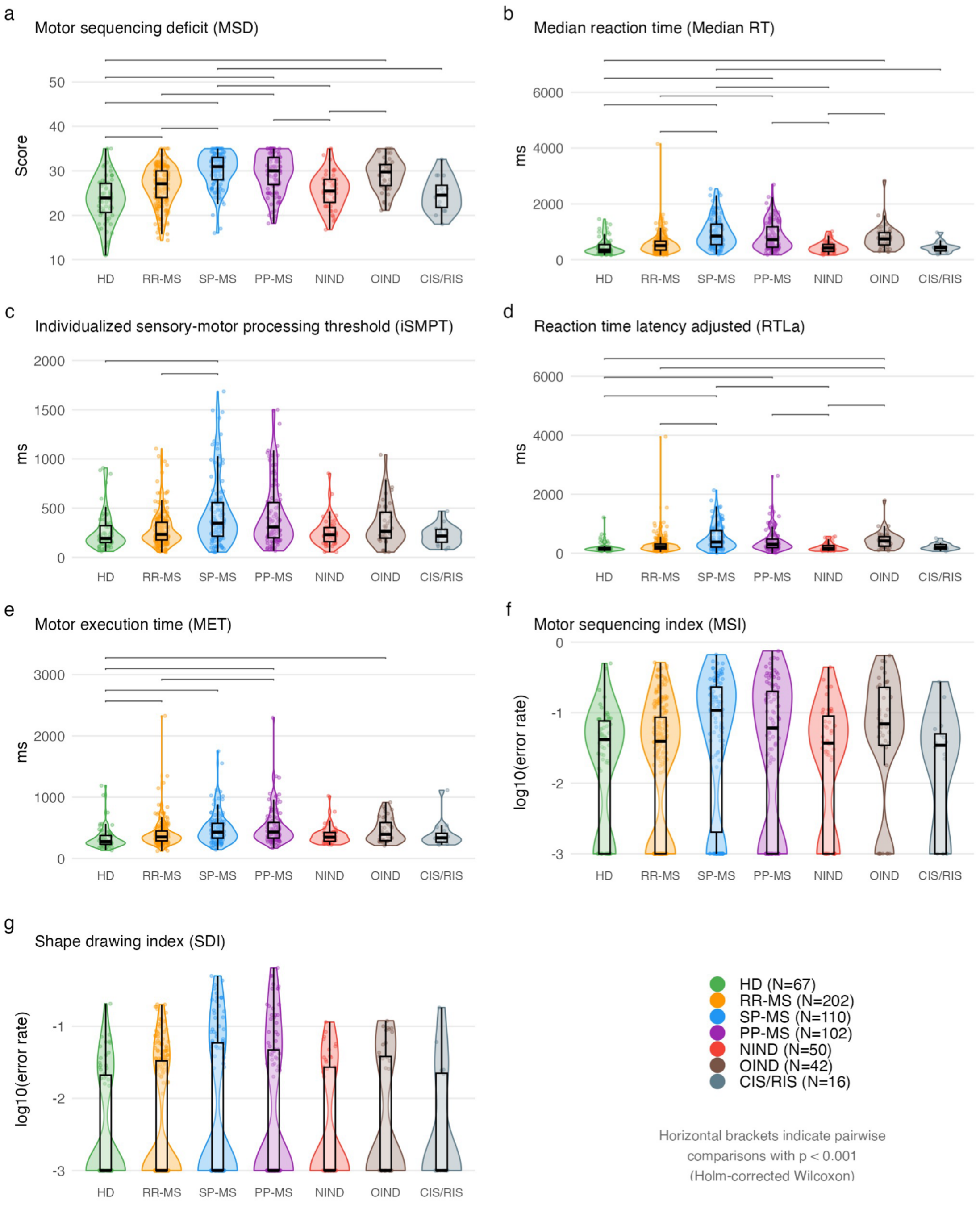


Supplementary Figure S6. Sensitivity analysis: discriminant validity across all diagnostic groups.

(a–g) Violin plots showing all seven biomarkers across all seven diagnostic groups (HD, green; RR-MS, orange; SP-MS, blue; PP-MS, purple; NIND, red; OIND, brown; CIS/RIS, gray).

Panels show: (a) MSD, (b) Median RT, (c) iSMPT, (d) RTLa, (e) MET, (f) MSI, (g) SDI. Horizontal brackets indicate significant pairwise comparisons. HD and NIND show comparable performance, OIND partially overlaps with MS, and CIS/RIS occupies an intermediate position. HD = healthy donors; RR-MS = relapsing-remitting MS; SP-MS = secondary-progressive MS; PP-MS = primary-progressive MS; NIND = non-inflammatory neurological disease; OIND = other inflammatory neurological disease; CIS/RIS = clinically/radiologically isolated syndrome; MSD = Motor Sequencing Deficit; Median RT = Median Reaction Time; iSMPT = individualized Sensory-Motor Processing Threshold; RTLa = Reaction Time Latency adjusted; MET = Motor Execution Time; MSI = Motor Sequencing Index; SDI = Shape Drawing Index.
